# Blood-derived microRNA signatures associated with hippocampal structure and atrophy rate: Findings from the Rhineland Study

**DOI:** 10.1101/2025.05.09.25327286

**Authors:** Konstantinos Melas, Valentina Talevi, Mohammed Aslam Imtiaz, Dennis Krüger, Tonatiuh Pena-Centeno, André Fischer, N. Ahmad Aziz, Monique MB Breteler

## Abstract

MicroRNAs have been linked to brain disorders, but their relations with hippocampal structure and atrophy remain unexplored. As the hippocampus is pivotal for cognition and dementia, understanding these relations and their specificity for the hippocampus would elucidate microRNA involvement in brain health and neurodegeneration. Here, using population-based data, we cross-sectionally and longitudinally examined the associations of blood-derived microRNAs with left and right hippocampal volume, hippocampal asymmetry, and total brain volume. Expression of microRNAs and their putative target genes was measured at study baseline in whole blood using RNA sequencing. Brain imaging measures were examined at baseline and re-examined 4.60 to 8.02 years later using 3T MRI. We investigated microRNA associations with imaging measures cross-sectionally using linear regression and longitudinally using linear mixed-effect models. Cross-sectionally, six microRNAs (miR-199a-3p, miR-199b-3p, miR-155-5p, miR-146a-5p, miR-6859-5p, miR-505-5p) were associated exclusively with left hippocampal volume. Longitudinally, another five microRNAs (miR-361-3p, miR-4473, miR-381-3p, miR-543, miR-370-3p) were associated with left hippocampal, right hippocampal, and total brain atrophy rates. Twenty-one microRNAs were exclusively associated with total brain atrophy rate. MicroRNAs identified in the cross-sectional analysis regulated target genes involved in brain development, memory, and synapse assembly. MicroRNAs from the longitudinal analysis regulated genes related to axonal and dendritic growth. Our findings suggest an asymmetric and specialized role of the cross-sectional microRNA signature during left hippocampal early-life development and a more universal role of the longitudinal signature during whole-brain aging or neurodegeneration. Importantly, some identified microRNAs have been previously linked to dementia and could be investigated as presymptomatic blood-based biomarkers.

## INTRODUCTION

MicroRNAs are small, non-coding, single-stranded RNAs with a critical role in the epigenetic regulation of neuronal development, function, and pathology (1,2). They regulate cellular functions by binding to and inhibiting target genes (3). As a single microRNA can bind to several target genes, its expression levels can reflect and regulate multiple (patho)physiological processes (4). Thus, they have been investigated for the diagnosis and therapy of neurodegenerative diseases, especially Alzheimer’s Disease (AD) (4–10).

However, in past studies, neurodegenerative diseases have mostly been dichotomized based on the presence or absence of a diagnosis. Although heuristically useful, this simplification disregards the heterogeneous nature of most such diseases as well as the potential interactions between microRNAs and the brain beyond disease, i.e., both during development and aging (11). To further elucidate the role of microRNAs in brain health and neurodegeneration, a detailed understanding of their association with key brain-related phenotypes is needed, coupled with a characterization of their biological functions. Moreover, identifying microRNAs associated with phenotypes typical of neurodegeneration in healthy, younger individuals could aid the development of microRNA biomarkers for the very early detection of neurodegenerative diseases.

The hippocampus is essential for long-term memory formation and spatial navigation (12). Medial temporal lobe and especially hippocampus-specific atrophy due to accumulation of neuropathology are considered characteristic of early AD and other dementias (13–16).

Notably, the left hippocampus tends to be smaller than the right (17) and this asymmetry increases in dementia (18), but there is little evidence for lateralized atrophy in normal aging (19,20). Additionally, a higher genetic risk of AD has been associated with a smaller hippocampus (21), which in turn has been associated with future cognitive decline (13,22) and worse memory performance, even in healthy, young individuals (23,24). Thus, the structure and atrophy patterns of the hippocampus in each hemisphere provide granular information on brain health and neurodegeneration. Previous studies have shown that blood-derived microRNA expression is related to brain atrophy in cohorts consisting of neurologic or psychiatric patients (25–31). However, the relationship of blood-derived microRNAs with hippocampal structure and atrophy in the general population remains unexplored. This relationship is particularly relevant for microRNAs suggested as biomarkers for the early detection of dementia (4), as it would support their involvement in preclinical neuropathology.

Here, we aimed to identify blood-derived microRNAs cross-sectionally and longitudinally related to hippocampal volume and left-to-right asymmetry, using data from the population-based Rhineland Study. To determine whether our findings were specific to the hippocampus, we also identified the microRNAs cross-sectionally and longitudinally related to total brain volume. Subsequently, we employed functional genomics to uncover genes and biological pathways regulated by the identified microRNAs. Moreover, we performed genome-wide association studies (GWAS) to detect microRNA expression quantitative trait loci (miR-eQTLs). Lastly, we leveraged these variants to assess potentially causal associations of microRNAs with imaging measures in a Mendelian randomization framework.

## METHODS

### Study Design

We based our analysis on data from the baseline and first follow-up examinations of the Rhineland Study, an ongoing prospective, population-based cohort study in Bonn, Germany. All residents of two pre-defined geographical areas of Bonn are invited to participate in the Rhineland Study. Inclusion criteria are age of 30 years or older and sufficient command of the German language to provide informed consent. Both at baseline and follow-up, each participant underwent a comprehensive 7-hour examination protocol.

### Standard protocol approvals, registrations, and participant consent

Approval to undertake the study was obtained from the ethics committee of the University of Bonn, Medical Faculty. The study is carried out in accordance with the recommendations of the International Conference on Harmonization Good Clinical Practice standards. We obtained written informed consent from all participants in accordance with the Declaration of Helsinki.

#### Analytical samples used for cross-sectional and longitudinal analyses

At study baseline, biomaterial was selected from the first 3000 participants of the Rhineland Study who provided blood samples for microRNA sequencing. Cross-sectional analyses were performed based on a subset of 2062 participants with complete baseline microRNA and hippocampal imaging data. For the longitudinal analyses, we related baseline microRNA data to baseline and follow-up hippocampal imaging data. Complete baseline microRNA and follow-up hippocampal imaging data were available in 1634 participants. The period between baseline and follow-up visits ranged from 4.60 to 8.02 years, with a median of 5.52 years and a total person-time of 12 750 years (**Supplementary Figure S1**).

#### Image acquisition and hippocampal segmentation

During both baseline and follow-up, eligible participants underwent the same one-hour imaging protocol using 3T MRI scanners (Siemens Prisma Magnetom, Erlangen, Germany) equipped with a 64-channel head-neck coil (32). T1-weighted images were used to obtain measurements of left and right hemisphere hippocampal volume, total brain volume, and estimated Total Intracranial Volume (eTIV) using the standard Freesurfer processing pipeline (version 6.0) (33,34). We evaluated total brain volume to assess whether our findings were specific to the hippocampus or reflected phenomena across the whole brain. Hippocampal and total brain volumes were normalized for eTIV using linear regression (see ‘Statistical analysis’). Hippocampal asymmetry was defined as (left hippocampal volume – right hippocampal volume)/(left hippocampal volume + right hippocampal volume).

#### Blood sample acquisition and storage

Blood samples were collected after overnight-fast between 7:00 to 9:45 in the morning. For microRNA and messenger RNA (mRNA) sequencing, samples were stored in PAXgene Blood RNA tubes (PreAnalytix/Qiagen) at -80° Celsius. Total RNA was isolated according to manufacturer’s instructions using the PAXgene Blood miRNA Kit, following the automated purification protocol (PreAnalytix/Qiagen) (35). For blood cell count measurements, blood samples were stored in ethylenediaminetetraacetic acid (EDTA) whole blood tubes.

#### MicroRNA and gene expression measurement

We performed the microRNA sequencing on the Illumina HiSeq 2000 platform measured over 44 plates and the mRNA sequencing on the NovaSeq6000 platform measured over 28 plates, as detailed before (35). MicroRNA and genes with overall mean expression greater than 15 reads and expressed in at least 5% of the participants were used for further analysis, leading to 415 unique microRNAs and 11 019 unique genes. Raw microRNA and gene counts were normalized and log-transformed before analysis (36).

#### General health variables and education

We evaluated the overall health and education of participants based on physician-diagnosed diseases (dementia, Parkinson’s disease, multiple sclerosis, hypertension, diabetes, stroke, coronary artery disease), smoking, and educational level. This information was based on self-reports, supplemented by clinical (for hypertension) and laboratory (for diabetes and smoking) measurements (**Supplementary Methods 1**).

#### Blood cell count measurement

Differential blood cell count measurements (erythrocytes, nucleated erythrocytes, leukocytes, and platelets), were performed at the Central Laboratory of the University Hospital Bonn, on a hematological analyzer (Sysmex XN9000).

#### Statistical analysis

We compared baseline characteristics between the analyzed subsets of our cohort using type III Analysis of Covariance adjusted for age and sex, except when examining age and sex themselves, where no adjustment was used.

Before further analyses, all numerical variables except age were z-standardized to enable comparison of effect sizes. We used multivariable linear regression to examine the cross-sectional association of microRNA expression (independent variable) with each imaging measure (dependent variable), adjusting for age, sex, and microRNA sequencing plate (batch). When assessing hippocampal or total brain volume as the dependent variables, we additionally adjusted for eTIV.

To examine whether microRNA expression at baseline was associated with longitudinal changes in imaging measures, we used linear mixed-effect models as follows:

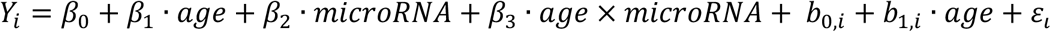

Here, *Y_i_* denotes the imaging measure at any time point for participant *i, β_0_* denotes the fixed mean intercept, *β_1_* denotes the fixed effect of age (i.e., the average change rate of the outcome per year across all individuals), *β_2_* is the fixed effect of baseline microRNA expression, *β_3_* is the fixed effect of the interaction between baseline microRNA expression and age, and *ε_i_* represents the residual error. For each participant, we also included a random intercept (*b_0,i_*) and a random slope for age (*b_1,i_*) to account for repeated measurements in time. The effect of baseline microRNA expression on the average change rate of imaging measures was thus given by the interaction term *β_3_*. The average change rate associated with one standard deviation (SD) increase of baseline microRNA expression was calculated as *β_1_* + *β_3_*, after converting coefficients to their original units *post-hoc* (i.e., mm^3^ for volumetric measures, ratio for hippocampal asymmetry). All available brain imaging data at any time point were included in the model and contributed towards the coefficients. As with the cross-sectional analysis, we adjusted the models for age, sex, microRNA batch, and in the case of hippocampal volume and total brain volume, for eTIV. We additionally calculated the proportion of the variance of change rates explained by statistically significant microRNAs. We created a base mixed-effect model with covariates and random effects as described above but not including any microRNAs, as well as a full model that additionally included all significant microRNAs and their interactions with age. We then calculated the proportion of the explained variance of the random slope as:

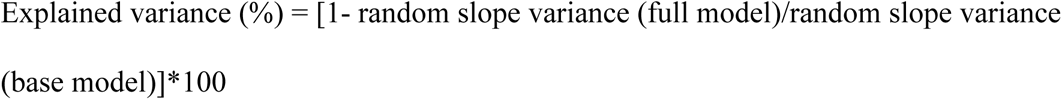

For both the cross-sectional and longitudinal analyses, we performed sensitivity analyses by additionally adjusting for baseline blood cell counts (leukocytes, erythrocytes, nucleated erythrocytes, and platelets). Additionally, we re-ran models after exclusion of participants who, at baseline, had a self-reported physician-diagnosed neurological disorder that could affect hippocampal volume, i.e., dementia (n=1), Parkinson’s disease (n=3), multiple sclerosis (n=10), hippocampal sclerosis (n=1).

To determine whether age and sex modified the association between microRNAs and imaging measures, we repeated the cross-sectional and longitudinal analyses in age and sex strata. Age strata were defined by the median age at baseline, i.e. age ≥ 54 years and age < 54 years.

The P-values of regression coefficients were corrected for multiple testing using the Benjamini - Hochberg false discovery rate (fdr) method (37) for the number of examined microRNAs (n=415). The statistical significance threshold was set at 0.05 (fdr ≤ 0.05). For microRNAs recently found to predict Mild Cognitive Impairment (MCI) and its conversion to AD (4), we also considered nominally significant P-values, i.e. without multiple testing adjustment.

All analyses were performed in R version 4.1.0.

#### MicroRNA expression in tissues and cells

To determine the tissue expression of microRNAs, we queried the miRNATissueAtlas2 (38). We first converted the expression of each microRNA across all tissues into a z-score. Next, for each tissue, we calculated the median of microRNA expression across donors and examined the tissues with the highest median expression. The miRNATissueAtlas2 also included the Tissue Specificity Index. This continuous metric ranges from 0 to 1. Values closer to 1 indicate that a microRNA is only expressed in a few or a single tissue.

To examine microRNA expression in cells we downloaded an atlas based on the Functional Annotation of Mammalian Genome (FANTOM5) project (39). We excluded cancer cell lines and cell lines treated with factors that would alter physiological microRNA expression. Then, we averaged microRNA expression across multiple samples and we z-transformed expression across cells for each microRNA to facilitate comparisons.

#### Functional genomics and pathway enrichment analysis

To identify potential target genes of microRNAs, we first obtained putative microRNA target genes from MirTarBase (40), TargetScan (41) and miRDB (42). We then used linear regression adjusting for age, sex, blood cell counts, microRNA sequencing batch, and gene sequencing batch to identify target genes negatively associated with their targeting microRNA. We used these genes to perform a pathway enrichment analysis in the Gene Ontology: Biological Processes database (43). We grouped similar terms with the *rrvgo* R Bioconductor package (44) and further clustered these terms in six broader categories using the simplifyEnrichment R Bioconductor package (45) (**Supplementary Methods 2**).

As an additional analysis, we filtered identified target genes for high expression in the hippocampus. We downloaded RNA consensus tissue gene data from The Human Protein Atlas v.23.0 (46) and set a cut-off of gene expression > 10 normalized Transcripts per Million in the hippocampal formation.

#### MicroRNA expression quantitative trait loci (miR-eQTLs) and Mendelian randomization analysis

We conducted a genome-wide miR-eQTL analysis adjusting for age, sex, microRNA batch, and the first ten genetic principal components (47). Genome-wide significance was defined as P-value ≤ 5×10^-8^. We then performed a two-sample Mendelian randomization analysis to examine whether the associations between microRNAs (exposure) and imaging measures (outcome) could be causal. We included as instrumental variables *cis*-miR-eQTLs, defined as those located within 1 MB of microRNA sequences and significant at the P-value ≤ 1×10^-5^ threshold, clumped based on linkage disequilibrium (r^2^ < 0.001 within a 10 Mb window) (48). Depending on whether microRNAs were identified in the cross-sectional or longitudinal analysis, we used summary statistics of published GWAS of left and right hippocampal volume (n=21 282 participants) (49) or hippocampal atrophy (n=15 640 participants) (50). For microRNAs associated with hippocampal volume cross-sectionally, we additionally examined for reverse causation by coding left or right hippocampal volume as the exposure and microRNA expression as the outcome (**Supplementary Methods 3**).

#### Data Availability

The Rhineland Study’s dataset is not publicly available because of data protection regulations. Access to data can be provided to scientists in accordance with the Rhineland Study’s Data Use and Access Policy. Requests for further information or to access the Rhineland Study’s dataset should be directed to.

## RESULTS

### Participant characteristics and descriptive statistics

Study participants were generally well-educated, with a relatively low prevalence of smoking, major cardiovascular, or major neurodegenerative diseases (**Table 1)**. At baseline, the right hippocampus was, on average, larger than the left (paired t-test P value < 10^-15^; **Table 1**). Right and left hippocampal volume were highly correlated (Pearson’s correlation coefficient: 0.90, 95% CI: 0.89 to 0.90). Bilateral hippocampal volume tended to decrease with age, especially in older participants (**Supplementary Figure S2**). On average, right hippocampal volume decreased faster (mean right hippocampal volume change per year: -24.75 mm^3^, 95% CI: -79.15 to 29.85 mm^3^; mean left hippocampal volume change per year: -22.52 mm^3^, 95% CI: -74.90 to 29.66 mm^3^; paired t-test P value: 9.12 ×10^-4^). The yearly change rates of left and right hippocampal volume were only moderately correlated (Pearson’s correlation coefficient: 0.56, 95% CI: 0.53 to 0.60).

**Table 1:** Participant characteristics at baseline for each subset of the study sample.

| **Variable** | **Subset 1:**<br>Baseline participants with<br>complete microRNA data,<sup>1</sup><br>N = 2928 | **Subset 2:**<br>Baseline participants with<br>complete baseline microRNA and<br>hippocampal MRI data,<sup>2</sup> N = 2062 | **P value,<sup>3</sup>**<br>Comparison<br>of Subsets 1<br>and 2 | **Subset 3:**<br>Baseline participants with complete<br>baseline microRNA and follow-up<br>hippocampal MRI data,<sup>4</sup> N = 1634 | **P value,<sup>3</sup>**<br>Comparison<br>of Subsets 2<br>and 3 |
| --- | --- | --- | --- | --- | --- |
| Sex, n(%) |  |  | 0.63 |  | 0.93 |
| Female | 1,627 (56%) | 1,160 (56%) |  | 917 (56%) |  |
| Male | 1,301 (44%) | 902 (44%) |  | 717 (44%) |  |
| Age (years), mean (SD) | 55.40 (14.35) | 54.48 (14.00) | **0.02** | 53.64 (13.13) | **0.06** |
| Age range (years) | 30.29 – 95.38 | 30.29 – 95.38 |  | 30.29 – 88.50 |  |
| Parkinson's disease, n(%) | 11 (0.4%) | 4 (0.2%) | 0.32 | 2 (0.1%) | 0.60 |
| Dementia, n(%) | 4 (0.1%) | 1 (<0.1%) | 0.38 | 1 (<0.1%) | 0.87 |
| Multiple Sclerosis, n(%) | 13 (0.4%) | 10 (0.5%) | 0.89 | 9 (0.6%) | 0.80 |
| Uncontrolled<br>hypertension, n(%) | 684 (23%) | 478 (23%) | 0.46 | 346 (21%) | 0.50 |
| Uncontrolled diabetes,<br>n(%) | 133 (4.5%) | 79 (3.8%) | 0.50 | 48 (2.9%) | 0.34 |
| Stroke, n(%) | 50 (1.7%) | 26 (1.3%) | 0.30 | 25 (1.5%) | 0.36 |
| Coronary artery disease | 129 (4.4%) | 59 (2.9%) | **0.04** | 46 (2.8%) | 0.45 |
| Educational level, n(%)<sup>5</sup> |  |  | 0.11 |  | 0.54 |
| low | 60 (2.1%) | 35 (1.7%) |  | 21 (1.3%) |  |
| middle | 1,276 (44%) | 850 (42%) |  | 658 (40%) |  |
| high | 1,565 (54%) | 1,160 (57%) |  | 946 (58%) |  |
| Current smoking, n(%) | 381 (13%) | 279 (14%) | 0.72 | 215 (13%) | 0.64 |
| Left Hippocampal Volume<br>(mm<sup>3</sup>), mean (SD) | - | 3.89×10<sup>3</sup> (0.46×10<sup>3</sup>) | - | 3.92×10<sup>3</sup> (0.45×10<sup>3</sup>) | 0.27 |
| Right Hippocampal<br>Volume (mm<sup>3</sup>), mean(SD) | - | 4.04×10<sup>3</sup> (0.50×10<sup>3</sup>) | - | 4.07×10<sup>3</sup> (0.49×10<sup>3</sup>) | 0.26 |
| Hippocampal asymmetry<br>index, mean(SD) | - | -0.02 (0.03) | - | -0.02 (0.03) | 0.32 |
| Total brain volume (mm<sup>3</sup>),<br>mean(SD) | - | 1.12×10<sup>6</sup> (0.12×10<sup>6</sup>) | - | 1.12×10<sup>6</sup> (0.12×10<sup>6</sup>) | 1.00 |
| Estimated total intracranial<br>volume (mm<sup>3</sup>), mean(SD) | - | 1.55×10<sup>6</sup> (0.15×10<sup>6</sup>) | - | 1.56×10<sup>6</sup> (0.16×10<sup>6</sup>) | 0.59 |

<sup>1</sup> The subsets used for functional genomics and miR-eQTL analyses were derived from Subset 1, after removing participants with missing gene expression data (n=565) or genetic data (n=472), respectively
<sup>2</sup> Data missing for total brain volume (n=17)
<sup>3</sup> Intergroup differences were tested using type III Analysis of Covariance
<sup>4</sup> Data missing for total brain volume (n=15)
<sup>5</sup> For educational level, analysis of covariance was performed for two groups: high education and middle-low education

### Cross-sectional association of microRNA expressions with imaging measures

Overall, significant baseline associations of microRNA expressions with hippocampal volume were more abundant and markedly stronger for the left hippocampus, while they differed from the associations with total brain volume. Specifically, higher expressions of miR-199a-3p, miR-199b-3p, miR-155-5p, miR-146a-5p, and miR-505-5p were significantly associated with larger left hippocampal volume, while higher expression of miR-6859-5p was significantly associated with smaller left hippocampal volume. While the same microRNAs were among the top results for right hippocampal volume, the effects were much weaker and no associations survived adjustment for multiple testing (**Figure 1**). As miR-199a-3p and miR-199b-3p have the same mature sequence (51), were very similarly distributed in our data (**Supplementary Figure S3**), and were similarly associated with hippocampal volume, we treated them as a single cluster (henceforth named miR-199a-3p/miR-199b-3p) for functional and genomics analyses Interestingly, higher expressions of three microRNAs, miR-125b-5p, miR-18a-5p, and miR- 26b-5p, recently found to predict conversion from early MCI to AD (4), were associated with larger left hippocampal volume at nominal significance.

**Figure 1:**
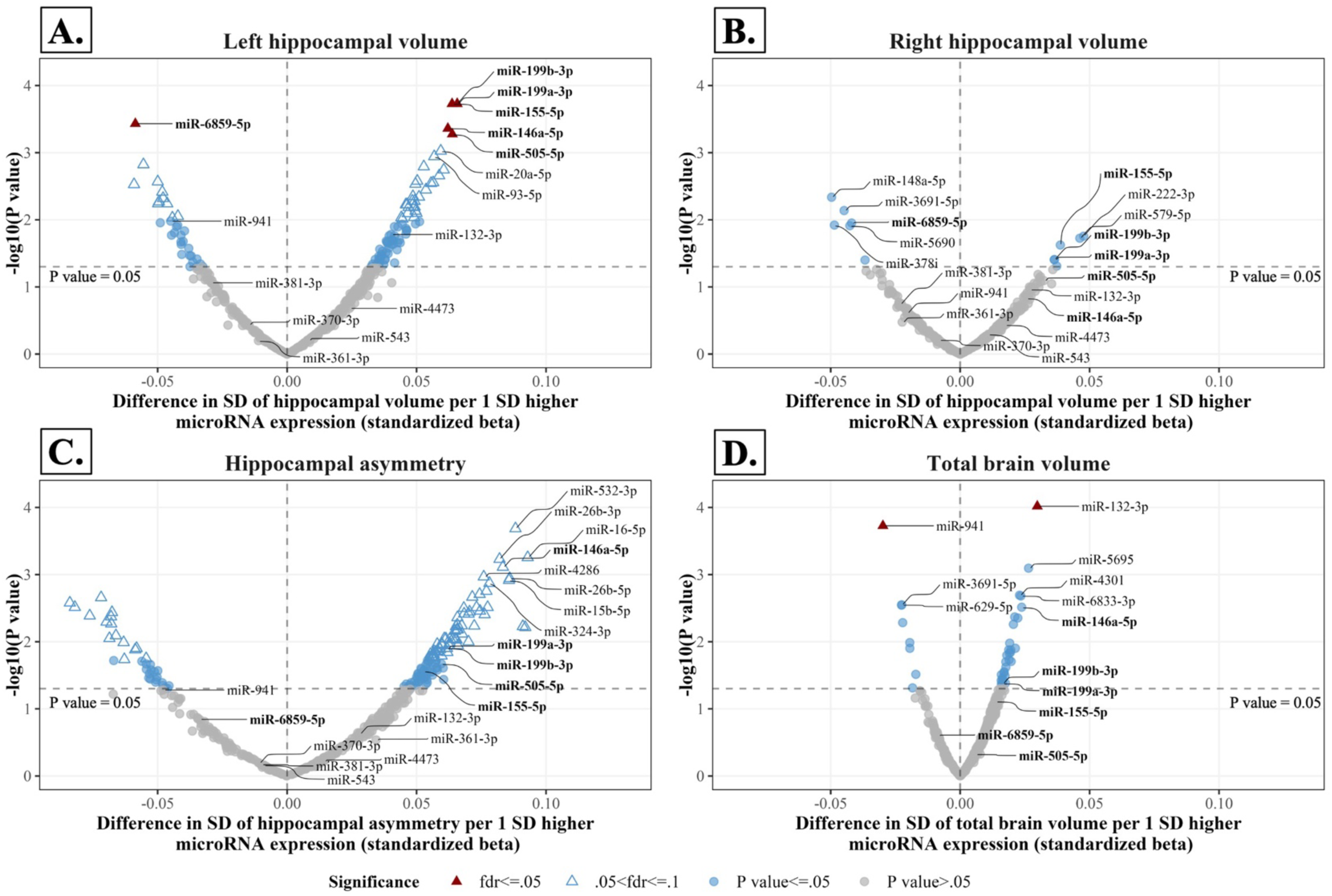
Cross-sectional associations of microRNAs with brain imaging measures. The volcano plots show the association of microRNAs with **A.** left hippocampal volume, **B.** right hippocampal volume, **C.** the hippocampal asymmetry index and **D.** total brain volume. MicroRNAs significantly associated with hippocampal volume after FDR correction are displayed in bold. Abbreviations: HV, Hippocampal Volume; SD, Standard Deviation; fdr, False Discovery Rate

The expressions of 77 microRNAs were associated with hippocampal asymmetry at borderline significance (fdr between 0.05 and 0.1), including most of the microRNAs associated with left hippocampal volume. However, none was significant at the fdr ≤ 0.05 threshold (**Figure 1C**). Significant associations with total brain volume were found for miR- 132-3p and miR-941 but for none of the microRNAs associated with left hippocampal volume (**Figure 1D**).

When additionally controlling for blood cell counts or removing participants with neurological diseases, associations of microRNAs with all MRI measures remained highly similar (**Supplementary Figure S4**). When stratifying by age, microRNA associations with left hippocampal volume remained similar and only slightly differed between younger (age < 54 years) and older (age ≥ 54 years) participants, but did not survive multiple testing adjustment **(Supplementary Figure S5).** When we stratified by sex, we observed little differences between men and women for left hippocampal volume, right hippocampal volume, and hippocampal asymmetry. However, four new microRNAs (miR-4473, miR-629- 5p, miR-4781-3p, and miR-330-5p) were significantly associated with total brain volume in men only **(Supplementary Figure S6).**

#### Association of baseline microRNA expressions with longitudinal change of imaging measures

Contrary to the cross-sectional analysis, we observed many similarities in the longitudinal associations of baseline microRNA expressions with the atrophy rates of the left hippocampus, right hippocampus, and total brain. Increased baseline expression of miR-361- 3p and miR-4473 was associated with significantly slower left hippocampal atrophy rates over time. For example, in the linear mixed-effect model for miR-361-3p, the average yearly change rate of left hippocampal volume was -17.86 mm^3^ (-0.46% per year compared to baseline). However, when baseline miR-361-3p expression was one SD higher, the average yearly change rate was modified to -15.98 mm^3^ (-0.41% per year compared to baseline), corresponding to a relative decrease of 10.52 % of yearly hippocampal atrophy. Similarly, increased baseline expressions of miR-381-3p, miR-370-3p, and miR-543 were associated with significantly slower right hippocampal atrophy rates over time (**Figure 2**). Notably, these three microRNAs are clustered in the genome at the 14q32 locus (51). The top microRNAs associated with left and right hippocampal atrophy rates were highly similar (**Figure 2**). Thus, we considered the abovementioned five microRNAs (miR-361-3p, miR- 4473, miR-381-3p, miR-370-3p, and miR-543) as associated with both left and right hippocampal atrophy. Importantly, when included together in a single model, expressions of these microRNAs jointly explained 27.95% and 18.07% of the variance of left and right hippocampal atrophy rates, respectively.

**Figure 2:**
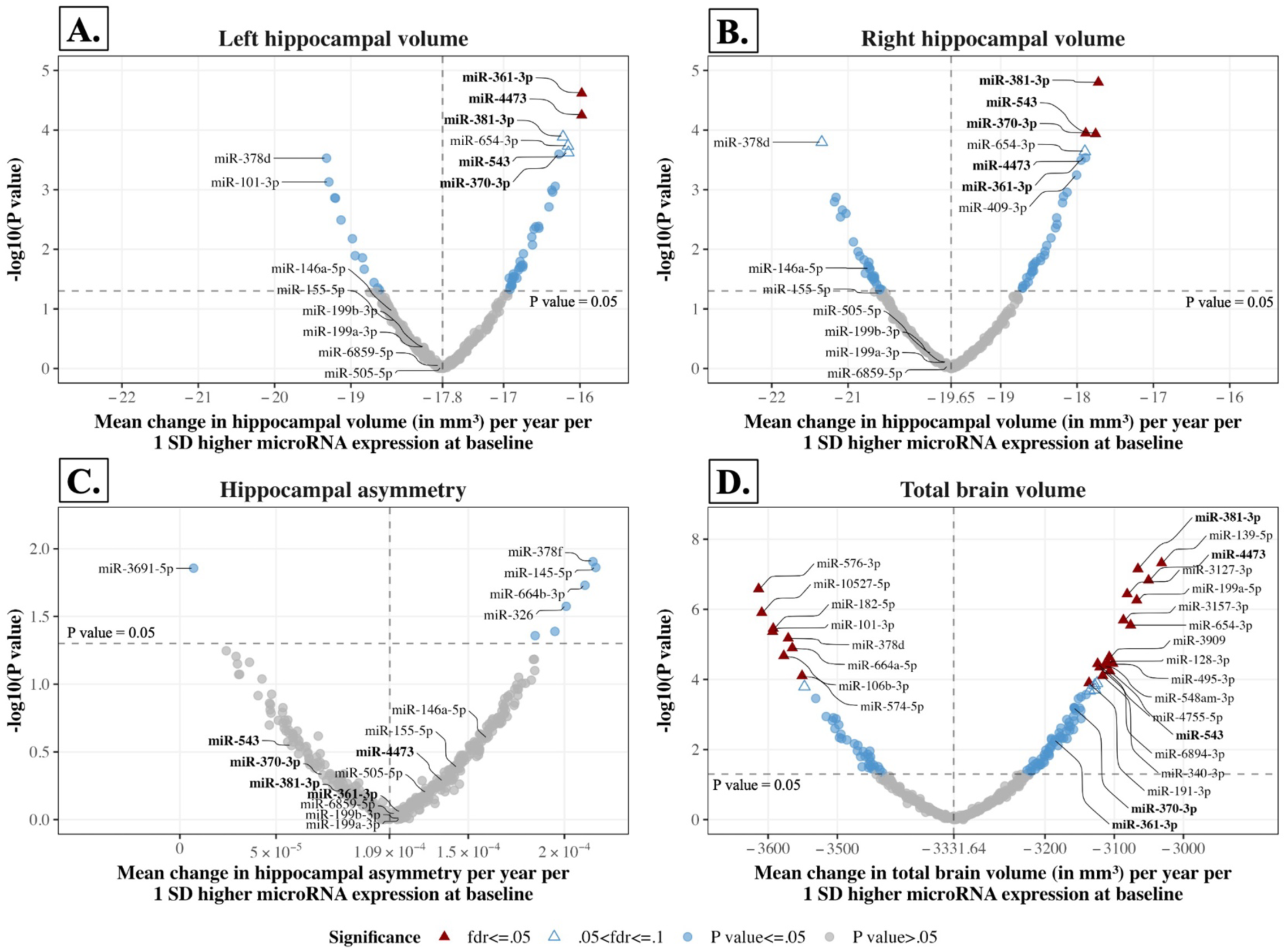
Longitudinal associations of baseline microRNAs with brain imaging measures. The volcano plots show the association of baseline microRNAs with the yearly change rates of **A.** left hippocampal volume, **B.** right hippocampal volume, **C.** hippocampal asymmetry, and **D.** total brain volume. MicroRNAs significantly associated with the yearly change of hippocampal volume after FDR correction are displayed in bold. The vertical dashed line indicates the yearly change rate of each measure when microRNA expression was equal to the population mean, averaged across all microRNAs. For microRNAs located at the right side of this line, higher baseline microRNA expression was associated with a slower rate of decline of imaging measures (e.g., slower hippocampal atrophy rate). To facilitate plotting, different axis scales have been used for hippocampal volume, asymmetry, and total brain volume. Abbreviations: SD, Standard Deviation; fdr, False Discovery Rate

When examining the longitudinal associations of microRNAs with hippocampal asymmetry, we found no significant results. Overall, longitudinal associations were stronger for total brain atrophy than hippocampal atrophy. Baseline expressions of 24 microRNAs were significantly associated with total brain atrophy rate over time. The five microRNAs associated with hippocampal atrophy rate were nominally associated with total brain atrophy rate, while miR-381-3p, miR-4473, and miR-543 survived adjustment for multiple testing (**Figure 2**).

Again, including blood cell counts in the model or removing participants with neurological diseases at baseline only minimally affected the longitudinal associations of microRNAs with the imaging measures (**Supplementary Figure S7**).

When stratifying by age, microRNA associations with hippocampal and total brain atrophy rates tended to be stronger in the younger group. The main exception was miR-361-3p, which had a much stronger association with left hippocampal atrophy rate in the older group.

Moreover, in the younger group, higher expressions of four different microRNAs were significantly associated with left and right hippocampal atrophy rates (left: miR-101-3p, miR- 106b-3p, right: miR-101-3p, miR-664a-5p, miR-3157-3p; **Supplementary Figure S8).** Sex stratification showed that microRNA associations with left and right hippocampal atrophy rates were generally similar in men and women. However, one microRNA, miR-125b-5p, was significantly associated with faster hippocampal atrophy rate in women only **(Supplementary Figure S9)**. This microRNA was among those recently found to predict conversion from early MCI to AD (4). Moreover, it was also associated with faster left and right hippocampal atrophy rates at a nominally significant threshold in our entire cohort.

#### MicroRNA expression in tissues and cells

Next, we examined the tissue expression of the microRNAs we identified in the cross-sectional and longitudinal analyses. We determined a high expression in brain tissues for that miR-505-5p and miR-6859-5p, from the cross-sectional analysis, as well as miR-361-3p, miR-4473, miR-381-3p, miR-543, and miR-370-3p from the longitudinal analysis. Moreover, miR-4473 was highly tissue-specific (Tissue Specificity Index: 0.83; **Figure 3**) and it highly expressed in neuronal cells (**Supplementary Figure S10**).

**Figure 3:**
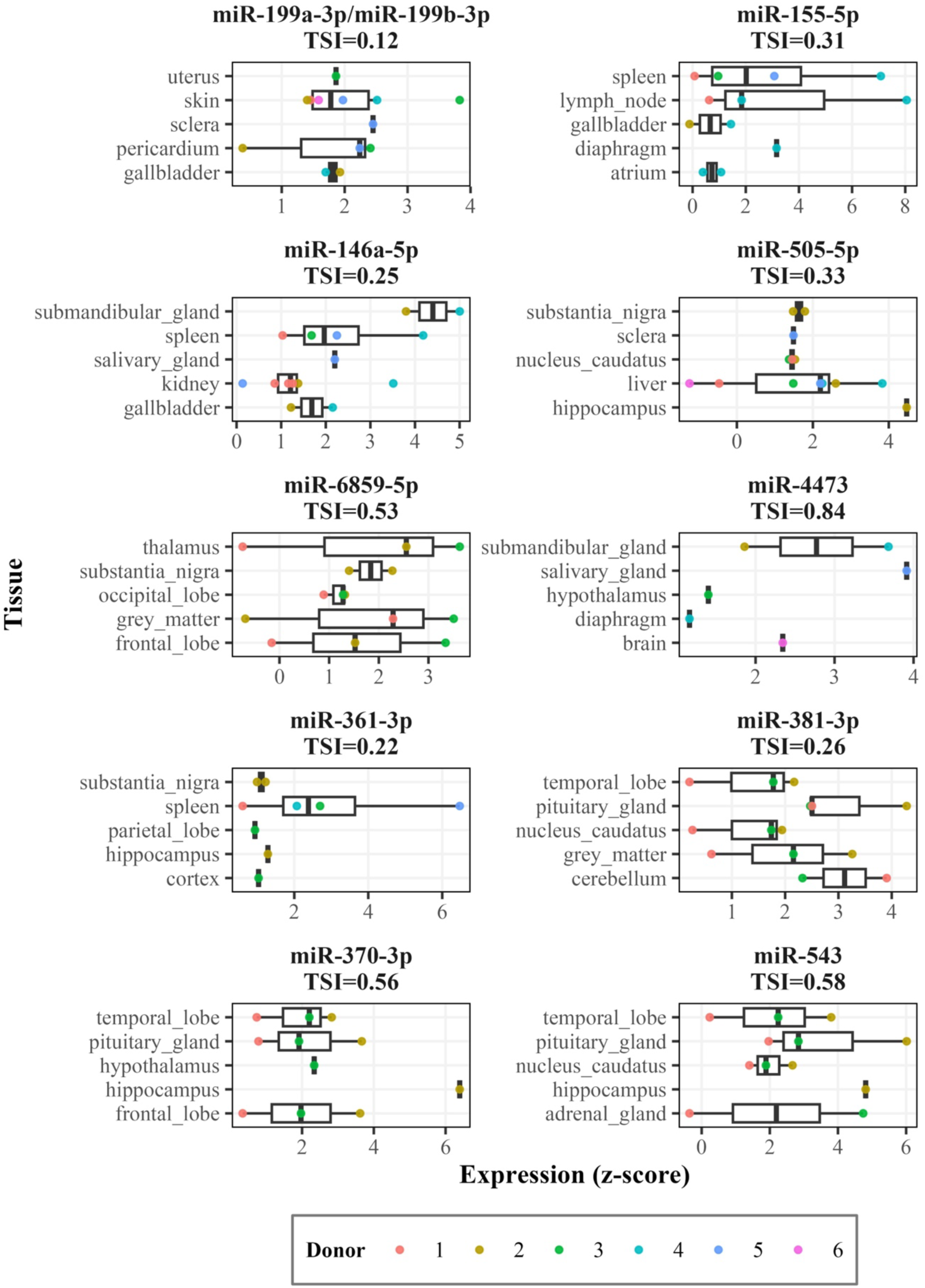
Tissue expression analysis of microRNAs associated with hippocampal volume cross-sectionally or longitudinally. Data was obtained from the miRNATissueAtlas (38), based on post-mortem samples taken from 6 human donors (here indicated by the colored dots). The boxplots show the top 5 tissues for each miRNA, based on median miRNA expression. Note that, to allow for better comparison of relative expression in tissues for each miRNA, the scale of the x-axis (expression z-score) varies. Abbreviations: TSI, Tissue Specificity Index

#### Functional genomics and pathway enrichment analysis

Functional genomics analysis determined that the microRNAs identified in the cross-sectional and the longitudinal analysis had 817 and 586 unique potential target genes, respectively **(Supplementary Figure S11A).** Generally, the identified microRNAs had relatively few common target genes (**Supplementary Figure S11B)**. Gene ontology enrichment of the target genes of each microRNA highlighted putative microRNA-regulated pathways. For example, miR-146a-5p targeted genes related to viral defense and inflammation, miR-6859-5p targeted genes related to cytoskeleton organization, and miR- 4473 targeted genes related to mitochondrial autophagy and cell growth. Overall, the majority of target genes of identified microRNAs were related to cellular development, signaling, and response to stimuli. MiR-155-5p, identified in the cross-sectional analysis, had the highest number of significantly enriched pathways (**Figures 4,5**).

**Figure 4:**
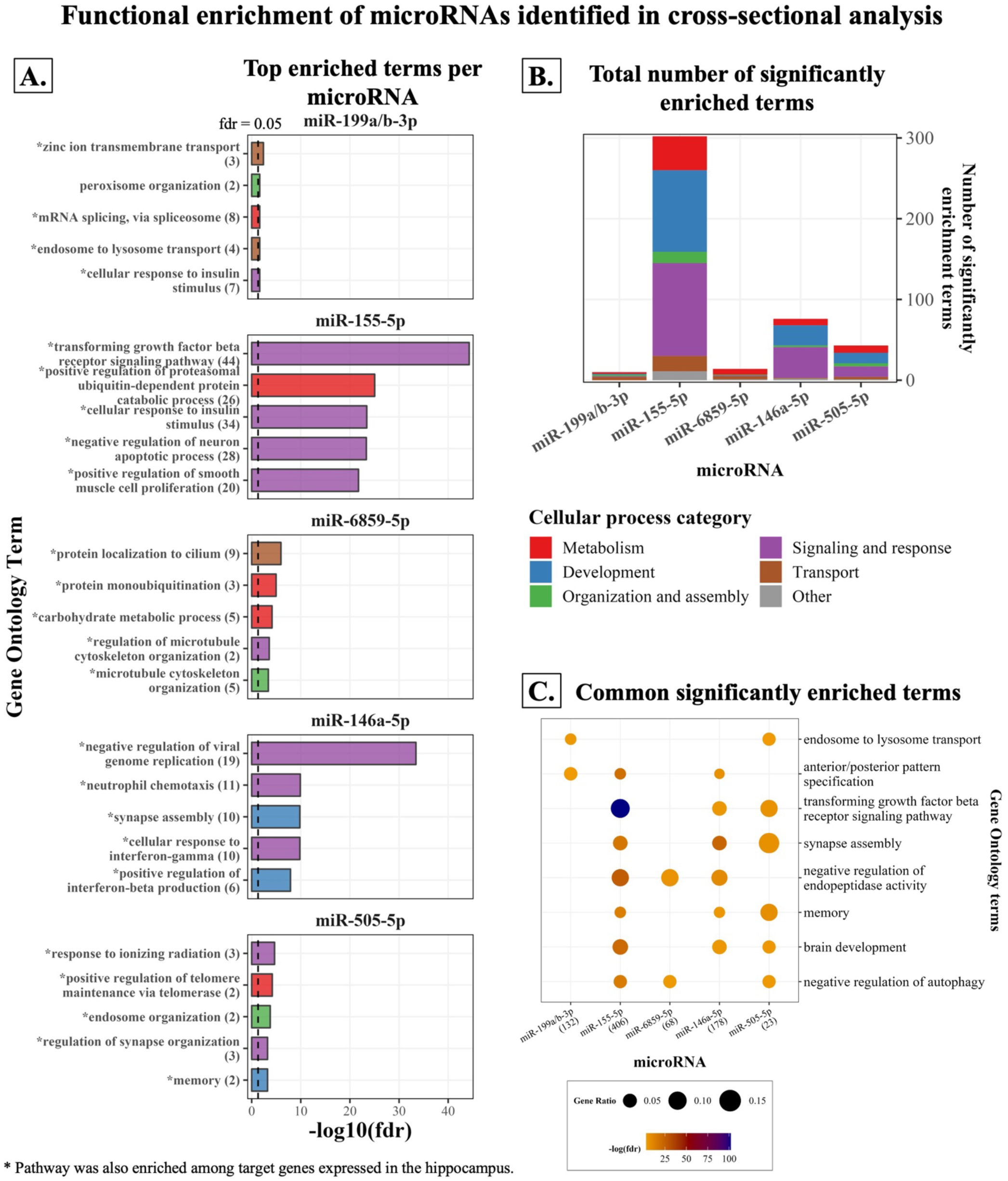
Results of *Gene ontology: Biological Process* enrichment analysis for target genes of microRNAs identified in the cross-sectional analysis. The top five significantly enriched pathways (lowest p-value) are shown in the bar plots in **A.**, with the number of target genes belonging to each pathway in parentheses. Pathways marked with an asterisk **(*)** were also enriched among target genes expressed in the hippocampus. Significantly enriched pathways for each microRNA were grouped in broad categories, indicated by bar colors. The total number of significantly enriched pathways for each microRNA is shown in **B.** Pathways enriched across the target genes of multiple (**≥** 3) microRNAs are shown in the bubble plot in **C.** Numbers in parentheses next to microRNA names indicate the total number of target genes of this microRNA that was included in *Gene Ontology*. P-values have been adjusted for multiple testing using the Benjamini-Hochberg false discovery rate method. Abbreviations: fdr, False Discovery Rate

**Figure 5:**
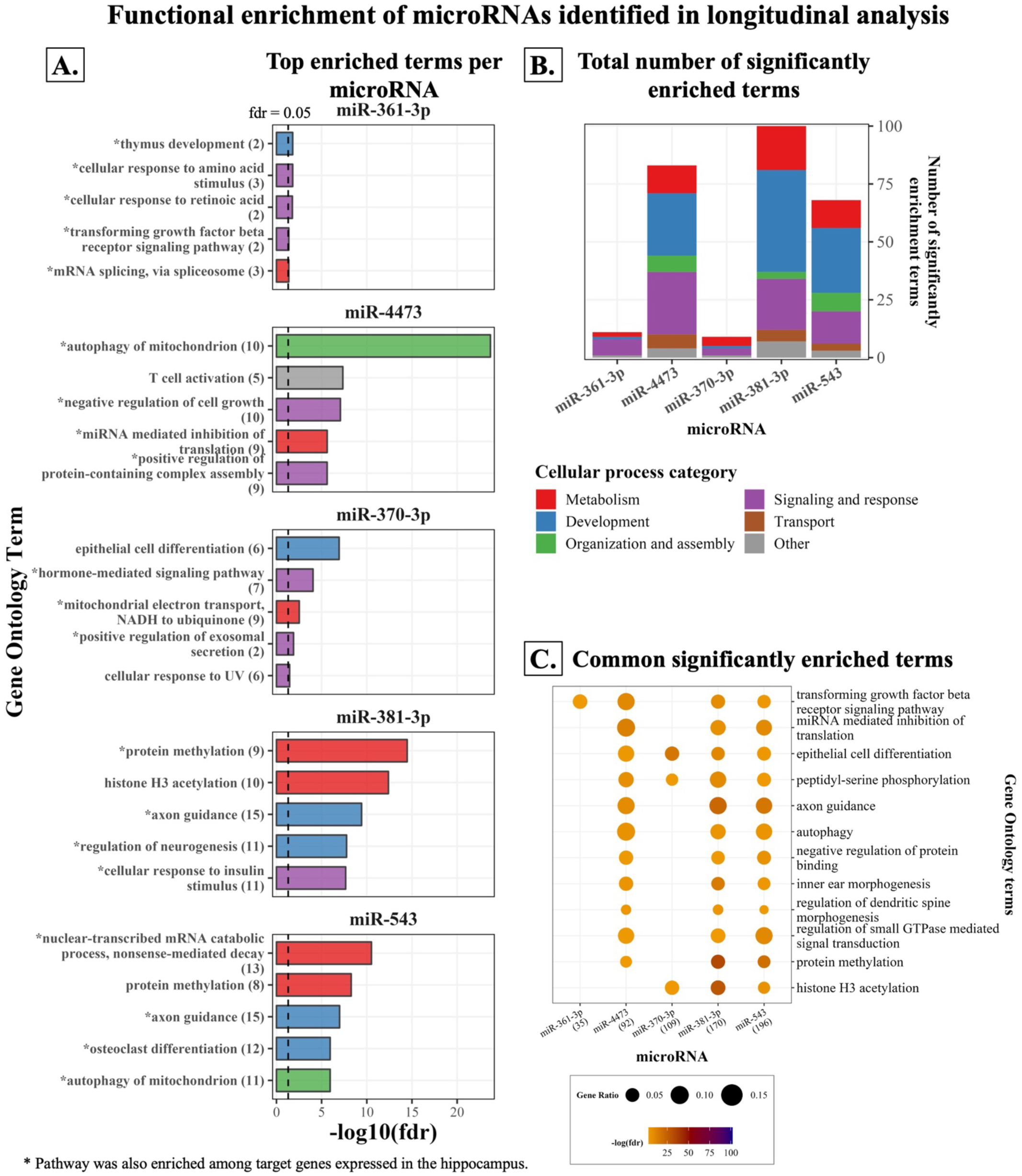
Results of *Gene ontology: Biological Process* enrichment analysis for target genes of microRNAs identified in the longitudinal analysis. The top five significantly enriched pathways (lowest p-value) are shown in the bar plots in **A.**, with the number of target genes belonging to each pathway in parentheses. Pathways marked with an asterisk **(*)** were also enriched among target genes expressed in the hippocampus. Significantly enriched pathways for each microRNA were grouped in broad categories, indicated by bar colors. The total number of significantly enriched pathways for each microRNA is shown in **B.** Pathways enriched across the target genes of multiple (**≥** 3) microRNAs are shown in the bubble plot **C.** Numbers in parentheses next to microRNA names indicate the total number of target genes of this microRNA that was included in *Gene Ontology*. P-values have been adjusted for multiple testing using the Benjamini-Hochberg false discovery rate method. Abbreviations: fdr, False Discovery Rate

Νotably, “transforming growth factor beta (TGF-β) receptor signaling” pathway was significantly enriched among targets of three microRNAs identified in the cross-sectional analysis (miR-155-5p, miR-146a-5p, miR-505-5p) and four microRNAs identified in the longitudinal analysis (miR-361-3p, miR-4473, miR-381-3p, and miR-543; **Figure 5C**).

Moreover, the “memory”, “brain development”, and “synapse assembly” pathways were each enriched for three of the microRNAs identified in the cross-sectional analysis (**Figure 4C)**.

The “axon guidance” and “dendritic spine morphogenesis” pathways were enriched for three of the microRNAs identified in the longitudinal analysis (**Figure 5C).** Lastly, miR-155-5p and miR-381-3p targeted genes related to amyloid-beta formation and clearance, including *PSEN1* for miR-155-5p.

We found high expressions in the hippocampus for 460 of the 822 (55.96%) target genes of microRNAs identified in the cross-sectional analysis and 329 of the 587 (56.05%) target genes of microRNAs identified in the longitudinal analysis. Most of the top enriched pathways identified in the main analysis were also enriched for hippocampus-specific genes (**Figures 4, 5** & **Supplementary Figure S12**).

#### miR-eQTL and Mendelian randomization analysis

Among the microRNAs from the longitudinal analysis, we identified in total 19 unique independent lead genome-wide significant (P value ≤ 5×10^-8^) *cis-*SNPs that were associated with the expressions of either miR-370-3p, miR-381-3p, miR-543, or miR-4473. Among the microRNAs identified in the cross-sectional analysis, 10 unique lead independent genome-wide significant *trans-*SNPs but no *cis*-SNPs were associated with the expressions of miR- 146a-5p or miR-6859-5p. We did not perform the miR-eQTL analysis for miR-505-5p and miR-361-3p, which are located on the X chromosome, for which no genotype data was available.

After clumping of all SNPs that reached suggestive significance (P value ≤ 1×10^-5^), 7 independent *cis*-miR-eQTLs remained as genetic proxies for miR-146a-5p from the cross-sectional analysis and miR-370-3p, miR-381-3p, miR-543, and miR-4473 from the longitudinal analysis. Using these SNPs as genetic instruments, we performed two-sample Mendelian randomization analyses to determine potential causal effects of miR-146a-5p on left hippocampal volume and right hippocampal volume, as well as miR-381-3p, miR-543, miR-370-3p, and miR-4473 on hippocampal atrophy rate. We found no evidence of a causal influence of these microRNAs on hippocampal volume. However, when assessing for reverse causation, we found evidence for a causal association of larger left and right hippocampal volume with lower miR-6859-5p expression. The F-statistic for all Mendelian randomization analyses ranged between 24.32 and 139.31, indicating a low chance of weak instrument bias.

## DISCUSSION

In the present study, we identified six microRNAs that were cross-sectionally associated with left hippocampal volume only. A different signature of five microRNAs was not only associated with hippocampal atrophy rate bilaterally, but also with total brain atrophy rate.

The difference in the results of the cross-sectional and longitudinal analyses suggests involvement of different microRNAs in hippocampal early-life development and aging. To further disentangle the biogenesis and function of identified microRNAs, we used genomics and transcriptomics data. Indeed, we found that many of them were genetically influenced and regulated pathways related to brain development, memory, synapse assembly, axon guidance, and dendrite morphogenesis.

Expressions of miR-199a-3p/miR-199b-3p, miR-155-5p, miR-146a-5p, miR-6859-5p, and miR-505-5p were cross-sectionally associated with left hippocampal volume, but not right hippocampal or total brain volumes. These distinct association patterns could indicate unique, lateralized epigenetic influences of microRNAs during hemispheric hippocampal development (52). Although no microRNAs were significantly associated with hippocampal asymmetry, this could be due to insufficient statistical power, as several were borderline significant. Our enrichment analysis supports a role in development for miR-155-5p, miR- 146a-5p, and miR-505-5p, which were enriched for brain development, axon guidance, and TFG-β signaling, a pathway crucial for brain development and function (53). For miR-199a- 3p/miR-199b-3p, involvement in hippocampal development is particularly supported by previous studies, as these microRNAs have been implicated in dendrite development (54), early life neurogenesis, and neuronal differentiation (55,56).

Beyond hippocampal development, the six microRNAs associated with left hippocampal volume cross-sectionally could be involved in processes affecting hippocampal volume later in life, such as aging or neurodegeneration. Indeed, miR-146a-5p and miR-155-5p are well known for their roles in neuroinflammation (57,58), hippocampal neurotoxicity (59,60), and AD pathology (61,62). Despite their putative neurotoxic role, these two microRNAs were associated with a larger hippocampus in our study. Similar discrepancies have been observed before and could be due to blood-derived microRNAs originating outside the brain, or due to altered cellular microRNA secretion by brain cells as a result of increased intracellular concentrations. For example, in line with our findings, two previous meta-analyses have found that miR-146a-5p is downregulated in the blood (63) and cerebrospinal fluid (5) of AD patients, but tends to be upregulated in their brain (63). In addition to AD, miR-146a-5p has been suggested as an early biomarker of age-related cognitive dysfunction (64). These past studies combined with our results suggest miR-146a-5p in particular as an excellent candidate for the early detection of AD.

Mendelian randomization analyses did not detect causal links between identified microRNAs and any of our cross-sectional or longitudinal hippocampus measures. However, the lack of significant results in Mendelian randomization analyses is not sufficient evidence to exclude causality (65). Our analysis was limited by having few to no identified genetic instruments for examined microRNAs, due to the relatively small sample used for microRNA-eQTL analysis. Moreover, investigations for reverse causation returned no significant results, except for miR-6859-5p. Mendelian randomization indicated that a larger hippocampus leads to lower expression of this microRNA. Our enrichment analysis suggests that this little-studied microRNA is involved in essential cellular functions like cytoskeleton organization. Thus, larger hippocampi with more or larger cells might require larger amounts of miR-6859-5p to maintain neurogenesis and axonogenesis (66), leading to its reduced secretion in the periphery (67). Further investigated, this finding would support miR-6859-5p as a robust indicator of hippocampal size, and therefore brain health.

In contrast to the cross-sectional analysis, a common signature consisting of miR-361-3p, miR-4473, miR-381-3p, miR-543, and miR-370-3p was similarly associated with the atrophy rates of the left hippocampus, right hippocampus, and whole brain, implying a universal role of these microRNAs during brain aging. The substantial proportion of the variance of hippocampal atrophy rates jointly explained by these microRNAs suggests that they might be effective as a biomarker panel for dementia. Despite the inconclusiveness of our Mendelian randomization analyses, past experimental studies indicate that these microRNAs might influence hippocampal volume. Specifically, miR-361-3p protected against AD-related neuronal injury and cognitive deficits (68). Additionally, miR-370-3p impaired hippocampal neurogenesis (69), miR-543 regulated neuronal differentiation and migration (70), while miR-381-3p attenuated neurotoxicity induced by Αβ (71) and ischemia (72). Importantly, in meta-analyses, miR-370-3p tended to be downregulated in the brain (63) and miR-381-3p was upregulated in the cerebrospinal fluid (5) of AD patients. Thus, these AD-related microRNAs were already associated with hippocampal atrophy rate in our relatively young and healthy cohort. Moreover, for miR-370-3p, this association was much stronger for the hippocampus than the whole brain. Given that early AD preferentially targets the hippocampus (16), this marks miR-370-3p as a good candidate for the early detection and prevention of AD. Conversely, miR-4473 was strongly associated both with hippocampal and total brain atrophy rates. In addition, it was cross-sectionally associated with total brain volume in men and was with high specificity expressed in the brain. Future studies could explore the potential of this little-studied microRNA as a novel, easily accessible marker of unhealthy brain aging.

A recent study identified miR-125b-5p, miR-18a-5p, and miR-26b-5p, as the best predictors of conversion from early MCI to AD (4). We found that these three microRNAs were cross-sectionally associated with left hippocampal volume at nominal significance. Additionally, miR-125b-5p was longitudinally associated with hippocampal atrophy rate, especially in women, and has been associated with working memory in a previous study by our group (47). These overlapping results suggest that these three microRNAs, and especially miR-125b-5p, are involved in early AD in a sex-specific manner and could be used for its detection.

Besides AD, the microRNAs we identified might mediate the effect of other factors influencing hippocampal volume, like exercise, cognitive stimulation, or inflammation (73–75). For example, miR-101-3p, miR-106b-3p, miR-664a-5p, and miR-3157-3p were only associated with hippocampal and total brain atrophy rates in participants younger than 54 years. These four microRNAs could be investigated for early therapeutical interventions against brain atrophy, potentially even mimicking the beneficial effects of lifestyle interventions on brain health.

Our study has considerable strengths. First, it was based on a large cohort with a wide age range, increasing precision and statistical power. Second, using next-generation sequencing allowed the untargeted measurement of microRNAs and mRNA transcripts. Lastly, we characterized the biogenesis and functions of identified microRNAs through a multi-omics analysis. Our study also has potential limitations. As is common for cohort studies, our single-center study recruited from a predefined geographic area, potentially limiting generalizability. Moreover, we used whole blood samples for RNA sequencing, so that measured microRNAs and mRNAs might have originated from blood cells. We used publicly available data to determine the origin of identified microRNAs and performed sensitivity analyses for blood cell composition. However, it was not possible to pinpoint their exact origin.

In conclusion, here we identified two different microRNA signatures associated with hippocampal volume and its rate of atrophy, suggesting distinct and asymmetrical epigenetic influences on the hippocampus throughout lifetime. Among these microRNAs, miR-146a-5p and miR-370 in particular have been suggested as biomarkers of AD and are good candidates for its presymptomatic detection. Moreover, our study identified novel associations of the little-studied miR-6859-5p and miR-4473 with hippocampal volume and atrophy rate. The effects of these microRNAs on brain function and their relation to neurodegeneration require further investigation. Lastly, our study identified genetic variants influencing the expression of many microRNAs, as well as thousands of microRNA-gene interactions and related biological pathways. These findings could be used by experimental studies to develop microRNA-based therapeutic strategies against age-related hippocampal atrophy or neurodegeneration.

## Data Availability

The Rhineland Study's dataset is not publicly available because of data protection regulations. Access to data can be provided to scientists in accordance with the Rhineland Study's Data Use and Access Policy. Requests for further information or to access the Rhineland Study's dataset should be directed to RS-DUAC[at]dzne.de.

## ACKNOWLEDGEMENTS

We wish to thank all participants of the Rhineland Study and the study personnel that was involved in data collection. We also thank the Data Management team of the Rhineland Study for their continued support in generating and accessing the data used in this project. Additionally, we would like to thank Dr. Dan Liu for her feedback on a pre-final version of the manuscript. This work was supported by the Federal Ministry of Education and Research grant [FKZ: 01KX2230] with the title "PreBeDem - Mit Prävention und Behandlung gegen Demenz” and the Helmholtz Association under the 2023 InnovationsPool. The Rhineland Study is funded by the German Center for Neurodegenerative Diseases (DZNE). Andre Fischer received funding from the DFG priority program 1738, SFB1286, SFB 1002, by Germany’s Excellence Strategy - EXC 2067/1 390729940, the ERA-Net Neuron project EPINEURODEVO and the JPND project EPI-3E.

## CONTRIBUTIONS

KM, VT, MAI, AF, NAA, and MMBB conceptualized the study and contributed methods. KM, VT, and MAI performed formal analysis and created visualizations. KM and MMBB wrote the original draft of the manuscript. KM, VT, MAI, DMK, TPC, AF, NAA, and MMBB reviewed and edited the manuscript. AF and MMBB acquired funding. MMBB and NAA supervised the project. MMBB supervises and administers the Rhineland Study. All authors read and approved the final manuscript.

