## Supplementary Materials for "Blood-derived microRNA signatures associated with hippocampal structure and atrophy rate: Findings from the Rhineland Study"


**Supplementary methods 1-3**
**Supplementary Figure S1:** Study analytical sample and subsets
**Supplementary Figure S2:** Longitudinal change of brain imaging measures
**Supplementary Figure S3:** Distribution of normalized counts of microRNAs associated with hippocampal volume
**Supplementary Figure S4:** Sensitivity analyses of the cross-sectional association of microRNAs with brain imaging measures
**Supplementary Figure S5:** Age stratification, cross-sectional analysis
**Supplementary Figure S6:** Sex stratification, cross-sectional analysis
**Supplementary Figure S7:** Sensitivity analyses of the longitudinal association of microRNAs with brain imaging measures
**Supplementary Figure S8:** Age stratification, longitudinal analysis
**Supplementary Figure S9:** Sex stratification, longitudinal analysis
**Supplementary Figure S10:** MicroRNA expression in cells.
**Supplementary Figure S11:** MicroRNA target genes identified through functional genomics
**Supplementary Figure S12:** Enrichment analysis of microRNA target genes expressed in the hippocampus.
**Supplementary Figure S13:** Clustering of *Gene Ontology: Biological Processes* with the *SimplifyEnrichment* package.
**References**

### Supplementary methods 1

#### Data collection for major cardiovascular and neurodegenerative diseases, education, and smoking

The existence of a physician diagnosis of dementia, Parkinson's disease, multiple sclerosis, untreated hypertension, untreated diabetes, stroke, and coronary artery disease was determined based on participant self-reports. Participants were classified as having untreated hypertension when they reported no regular use of antihypertensive medication and either reported having a diagnosis of hypertension, had a mean systolic blood pressure exceeding 140 mmHg, or had a mean diastolic blood pressure exceeding 90 mmHg. Systolic and diastolic blood pressure were measured in a sitting position, using an oscillometric blood pressure device (Omron 705 IT) (1). The measurements were performed thrice, separated by ten minutes, by experienced study technicians while participants were sitting in a resting chair in a quiet environment. The mean of the second and third measurements was used for analyses. Participants were classified as having untreated diabetes when they reported no use of antidiabetic medication and either reported having a diagnosis of diabetes, had a glycated hemoglobin percentage exceeding 6.5%, or had a glucose exceeding126 mg/dL in fasting morning blood.

Information on education and current smoking was based on self-reports. Education was classified as "low" (early childhood education up to lower secondary education), "middle" (secondary education up to Bachelor's or equivalent level), or "high" (Master's or equivalent level up to Doctoral or equivalent level). Participants with missing self-reported data on smoking and with a cotinine level exceeding the non-smoker sample-defined 97.5 percentile were classified as current smokers.

### Supplementary methods 2

#### Functional genomics analysis

For microRNAs associated with imaging measures, we employed a functional genomics analysis to identify potential microRNA target genes. First, we used the *multimir* R package (v.1.12.0) (2) to obtain a list of putative target genes for each microRNA, from 3 online databases:  the MirTarBase (3) database was inquired for experimentally validated microRNA-target gene interactions, and the TargetScan (4) and miRDB (5,6) databases were inquired for predicted microRNA-target gene interactions. Each microRNA-target gene interaction obtained through the *multimir* package is accompanied by a prediction score, which is a measure of prediction confidence and is provided by the inquired databases. To identify all putative microRNA-target genes, we included all predicted target genes, regardless of the prediction score. Moreover, we employed the union of the target genes obtained from the 3 databases to minimize false negatives (7). Subsequently, we determined the association between microRNA expression and expression of their putative target genes using individual-level microRNA and gene expression data, available from 2363 participants of our study. We ran separate regression models, for each microRNA (independent variable) and each of its putative target genes (dependent variable), adjusting for age, sex, blood cell counts, microRNA sequencing batch, and gene seque4ncing batch. Lastly, we filtered for target genes that were negatively associated with their targeting microRNA (linear regression beta coefficient ≤ 0 and P value ≤ 0.05). P-values for this analysis were not adjusted for multiple testing, as the analysis was performed only for genes putatively targeted by microRNAs, providing an *a priori* hypothesis that they might be associated.

**Pathway enrichment analysis**

For the microRNA target genes identified in the previous steps, we performed a pathway enrichment analysis using the "*clusterProfiler*" (v. 3.18.1) R Bioconductor package (8,9). We applied an overrepresentation analysis in enrichment terms obtained from the Gene Ontology: Biological Processes (Gene Ontology) database (10,11). The universe (background genes) of the overrepresentation analysis was set to the genes measured in our study. Statistical significance was determined after adjusting for multiple testing with the false discovery rate (fdr) method and at a threshold of fdr ≤ 0.05.

Subsequently, redundant Gene Ontology terms were grouped using the rrvgo R Bioconductor package (v. 1.2.0) with default parameters (12). Specifically, a semantic similarity matrix of terms was constructed and similar terms were grouped, selecting the largest term (as defined by the number of genes in each enriched term) to represent each group. A relatively small similarity threshold of 0.6 was selected, so that only highly redundant terms were grouped. The P values of the terms within each group were pooled using the Fisher method and multiple testing adjustment with the BH method was performed for the pooled P values. To determine the broader cellular processes that the enriched terms belong to, we further clustered them into six large categories. This was done using the *simplifyEnrichment* R Bioconductor package (13). Specifically, we created a similarity matrix of terms based on gene overlap. We then used the *simplifyGo* function with default parameters to cluster terms based on the similarity matrix, setting the minimal number of terms within a cluster to 7. Resulting clusters were then named based on the most common terms, after visual inspection of word clouds (**Supplementary Figure S13**).

### Supplementary methods 3

**miR-eQTL analysis**

To identify microRNA expression quantitative trait loci (miR-eQTLs), we conducted a genome-wide miR-eQTL analysis on 2456 participants, for which both genetic and microRNA expression data was available. We evaluated the association between each SNP (independent variable) and expression levels of each microRNA (dependent variable) using multivariable linear regression, adjusted for age, sex, microRNA batch, and the first ten genetic principal components. *Cis*-SNPs were defined as those located within 1 Mb (1 Mb before the start or 1 Mb after the end) of the mature microRNA genomic sequence (14). The genome-wide significance level for *cis* miR-eQTLs was set at P value ≤ $5\times10^{-8}$. We used the Functional Mapping and Annotation (FUMA) for GWAS platform (15) to define genomic risk loci, by clumping SNPs in linkage disequilibrium at $r^2 > 0.6$. Significant SNPs in relatively high linkage disequilibrium at $r^2 < 0.6$ were defined as independent significant SNPs, while significant SNPs in approximate linkage disequilibrium at $r^2 < 0.1$ were defined as lead SNPs. We mapped genes to lead SNPs with positional mapping.

**Two-sample Mendelian Randomization**

We employed a Two-sample Mendelian Randomization analysis to examine whether the cross-sectional or longitudinal associations between microRNAs and imaging measures could be causal, using the TwoSampleMR R package (16). We included as instrumental variables *cis*-SNPs that were significant at the suggestive P value $\leq 1\times10^{-5}$ significance level in the miR-eQTL analysis (14) and microRNA expression as the exposure. Depending on whether microRNAs were identified in the cross-sectional or longitudinal analysis, we used as outcome hippocampal volume or atrophy based on published summary statistics from recent genome-wide association studies (GWAS) (17,18). Before the analysis, we clumped SNPs in linkage disequilibrium, defined as $r^2 < 0.001$ within a 10 Mb window. We performed the Mendelian Randomization analysis using the Wald ratio test when only one SNP instrumental variable was available, the inverse variance weighted method, when two SNP instrumental variables were available, and the inverse variance weighted, MR Egger, simple mode, weighted mode, and weighted median methods, when three or more SNP instrumental variables were available. We additionally examined for inverse causation by performing the same analysis, but this time, coding hippocampal volume or atrophy as the exposure and microRNA expression as the outcome. To assess the risk of weak instrument bias, we calculated the F-statistic for the selected SNP instrumental variables.

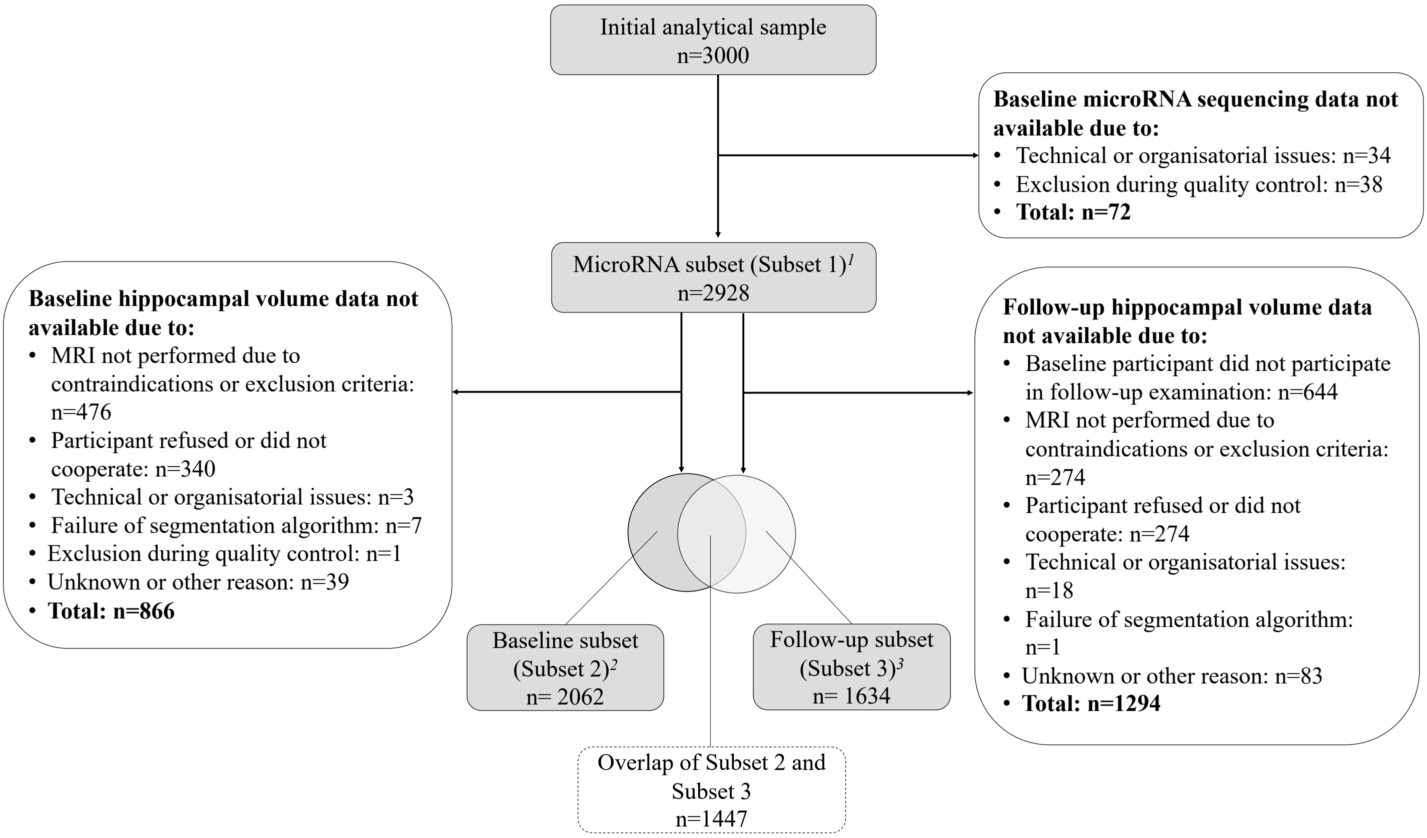


**Supplementary Figure S1:** Study analytical sample and subsets

[1] The subsets used for functional genomics and miR-eQTL analyses were derived from Subset 1, after removing participants with missing gene expression data (n=565) or genetic data (n=472), respectively.

[2] Data additionally missing for total brain volume: n=17.

[3] Data additionally missing for total brain volume: n=15.

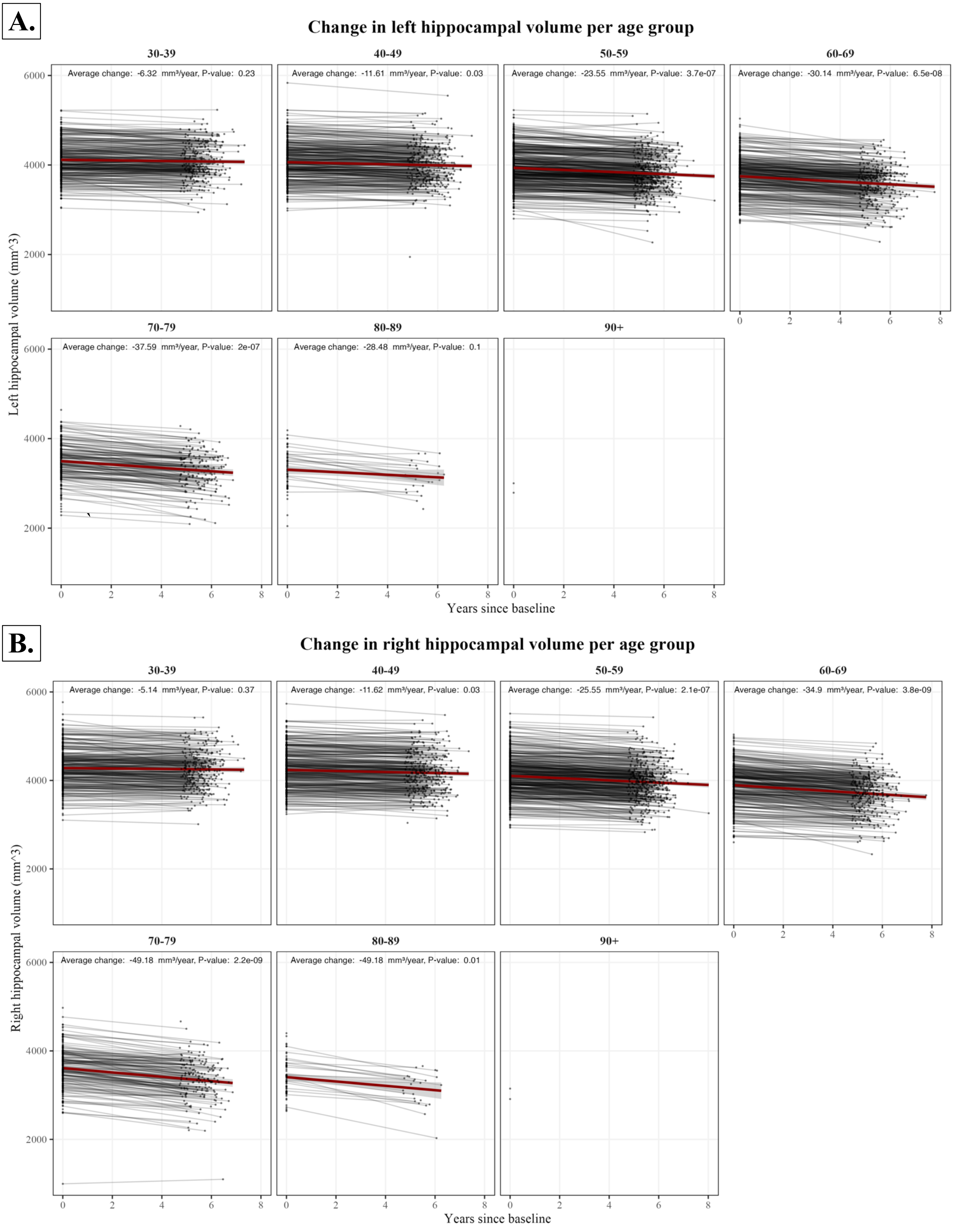


**Supplementary Figure S2:** Longitudinal change of brain imaging measures

**C.**

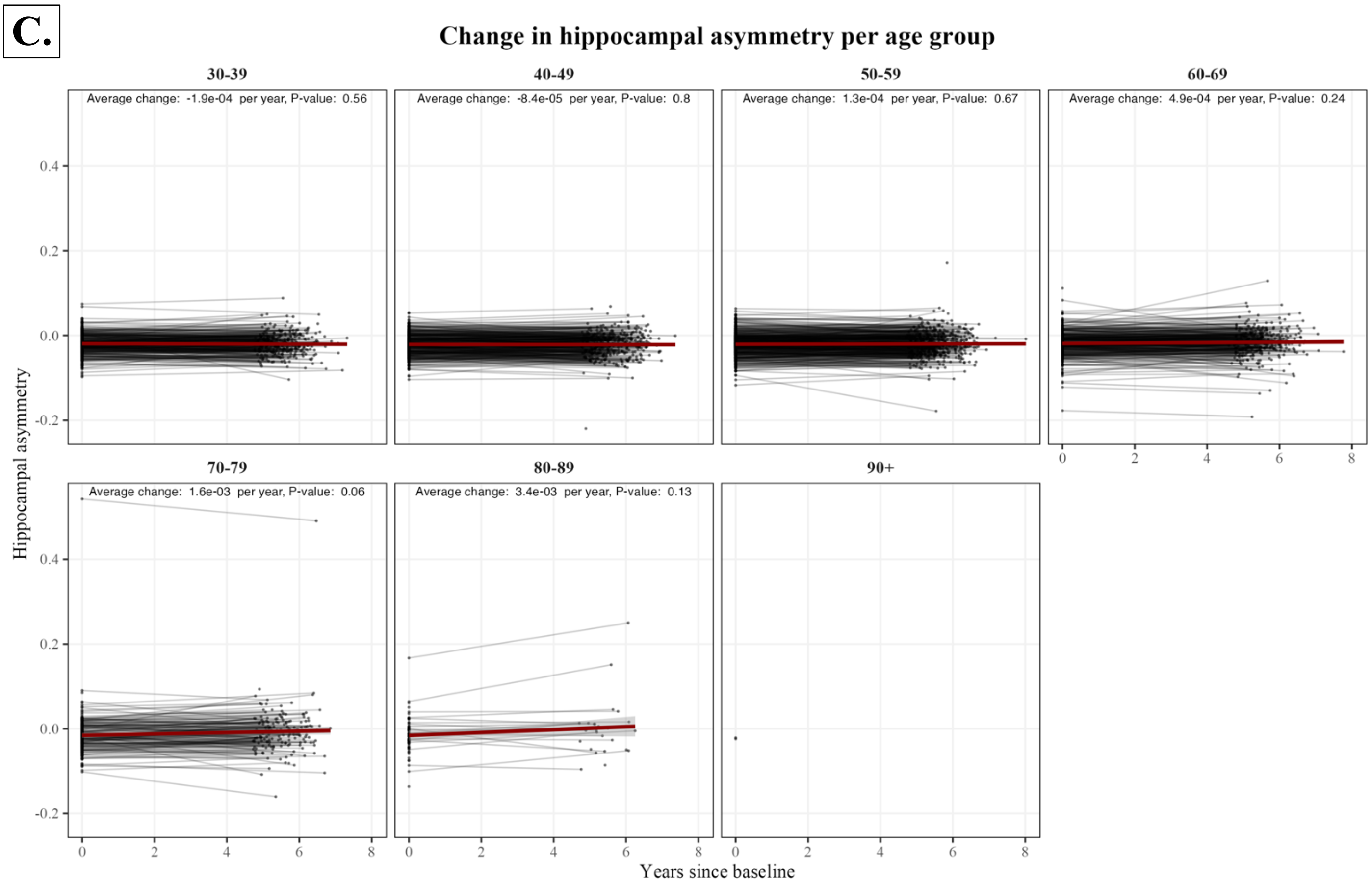


**D.**

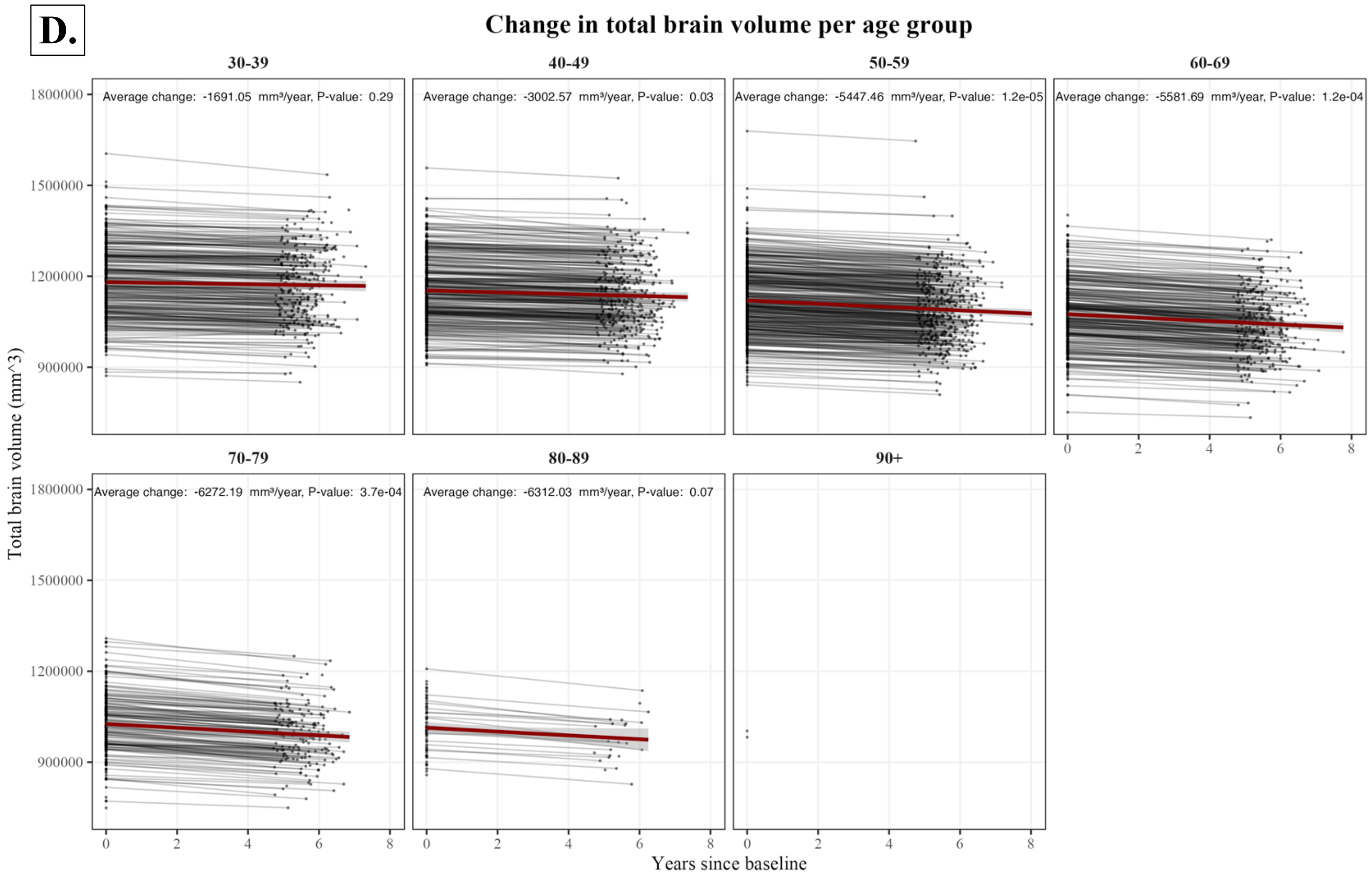


**Supplementary Figure S2 (cont.):** Longitudinal change of brain imaging measures
Each participant is represented by a single line connecting brain imaging measures as evaluated at baseline and the second round of examinations: left hippocampal volume (**A**), right hippocampal volume (**B**), hippocampal asymmetry (**C**), and total brain volume (**D**). Each dots represent one measurement. Participants have been stratified by age at baseline. The red line indicates the average change of participants in each stratum and the shaded area the corresponding 95% confidence interval. Average change rates and corresponding p-values have been calculated using linear regression

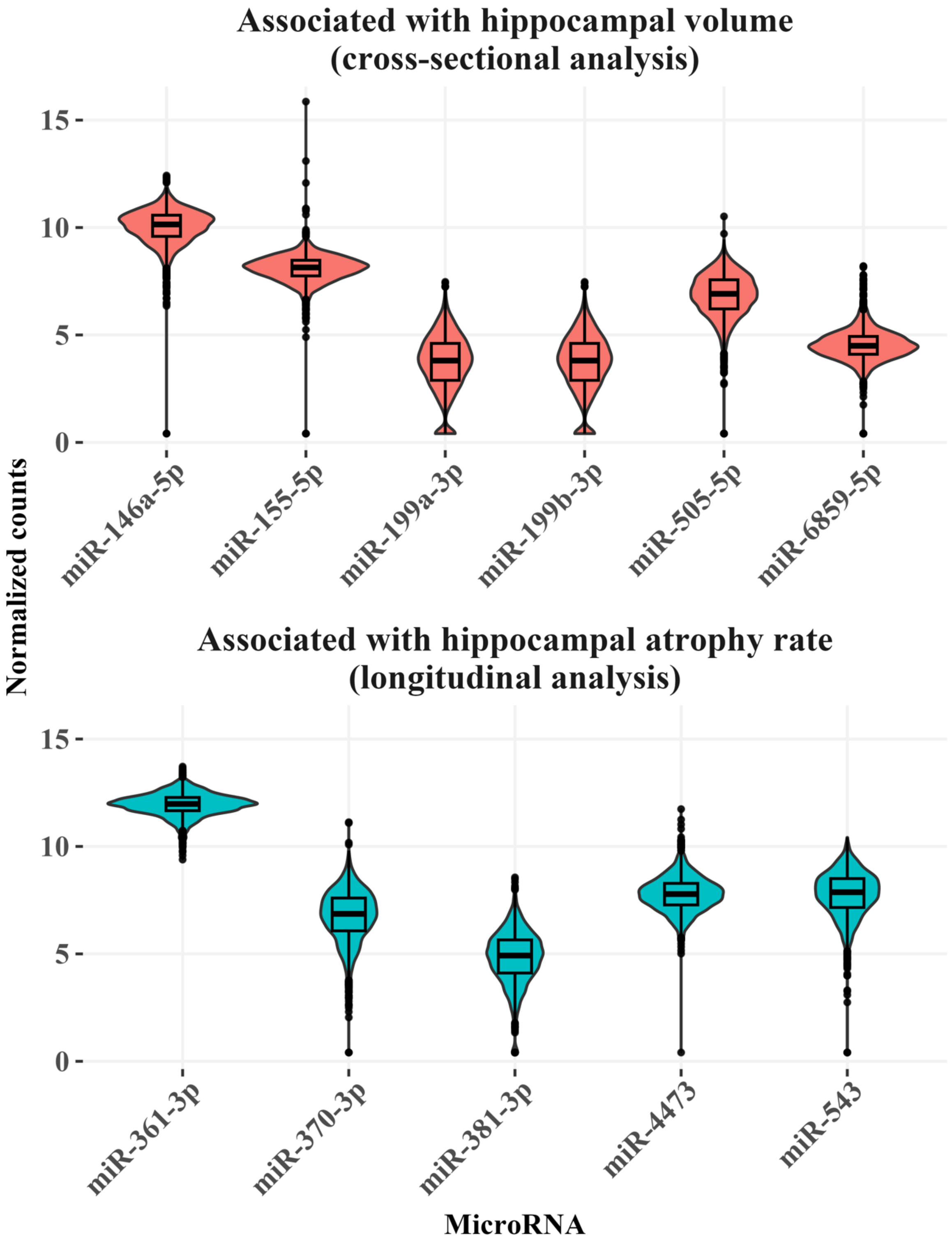


**Supplementary Figure S3:** Distribution of normalized counts of microRNAs associated with hippocampal volume and atrophy rate

The violin boxplots show the normalized and log-transformed counts for microRNAs identified in the cross-sectional or longitudinal analysis. The plots were created using all available microRNA data, regardless if brain imaging data was available.

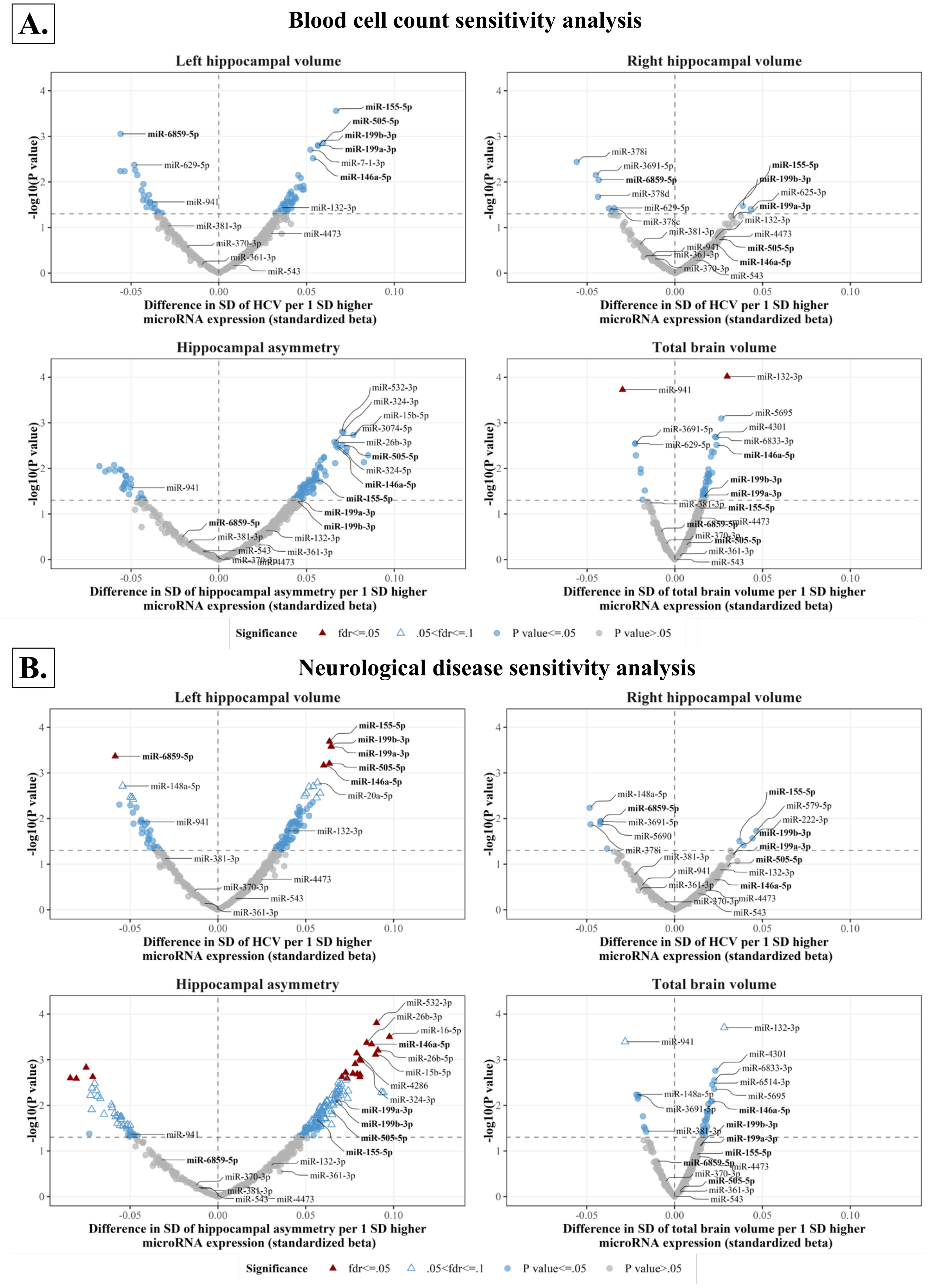


**Supplementary Figure S4:** Sensitivity analyses of the cross-sectional association of microRNAs with brain imaging measures.
The volcano plots show the association of microRNAs with brain imaging measures when adjusting for baseline blood cell counts in addition to age, sex and technical variables (**A.**) and after removing participants diagnosed with dementia, Parkinson's disease, multiple sclerosis and hippocampal sclerosis (**B.**). MicroRNAs annotated with bold letters were significantly associated with HCV in the main analysis
Abbreviations: SD, Standard Deviation; fdr, false discovery rate; HCV, Hippocampal Volume

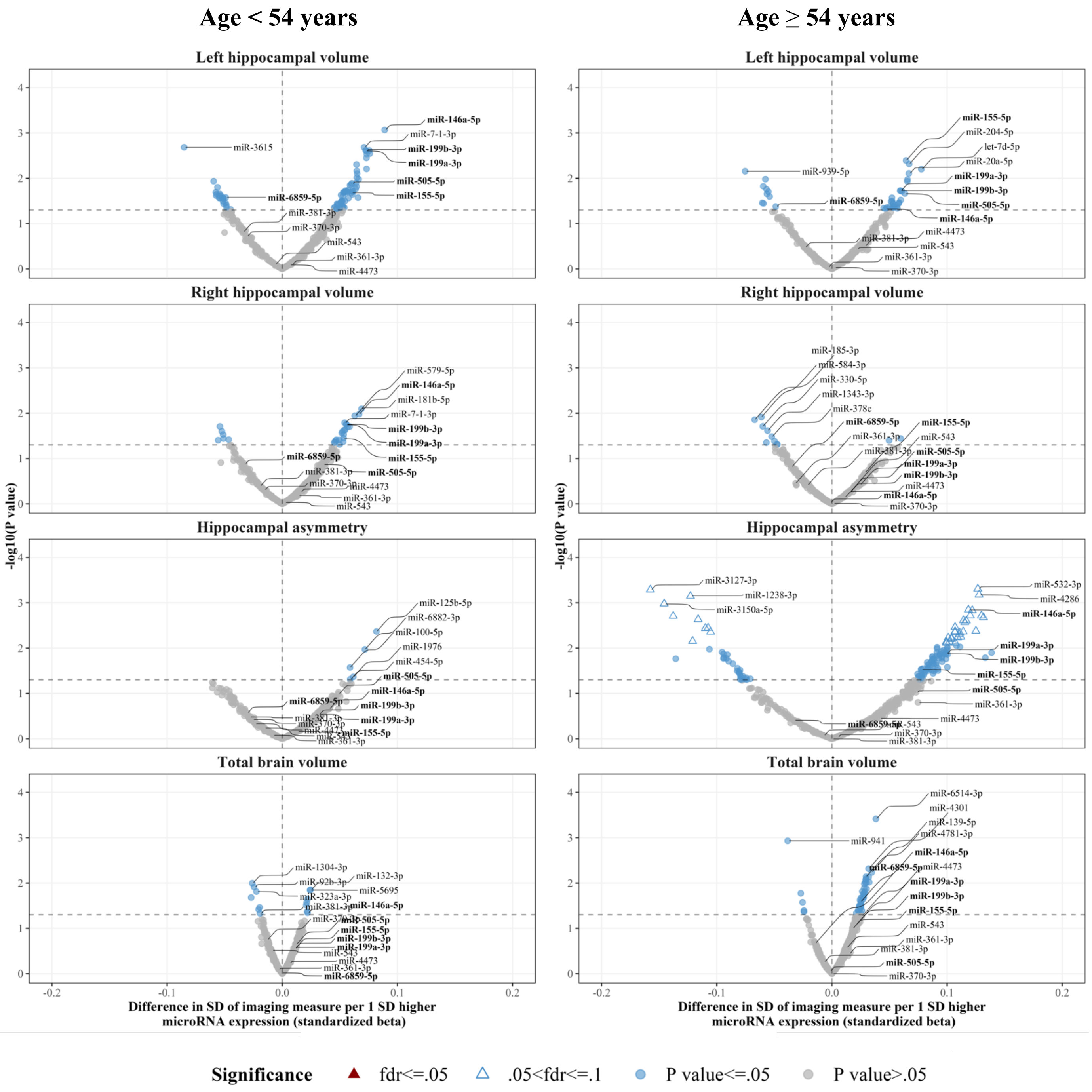


**Supplementary Figure S5:** Age stratification, cross-sectional analysis
The volcano plots show the association of microRNAs with brain imaging measures in younger (age < 54 years) and older (age ≥ 54 years) participants. MicroRNAs significantly associated with HCV in the main analysis which included all participants have been annotated with bold letters.
Abbreviations: HCV. Hippocampal Volume; SD, Standard Deviation; fdr, False Discovery Rate

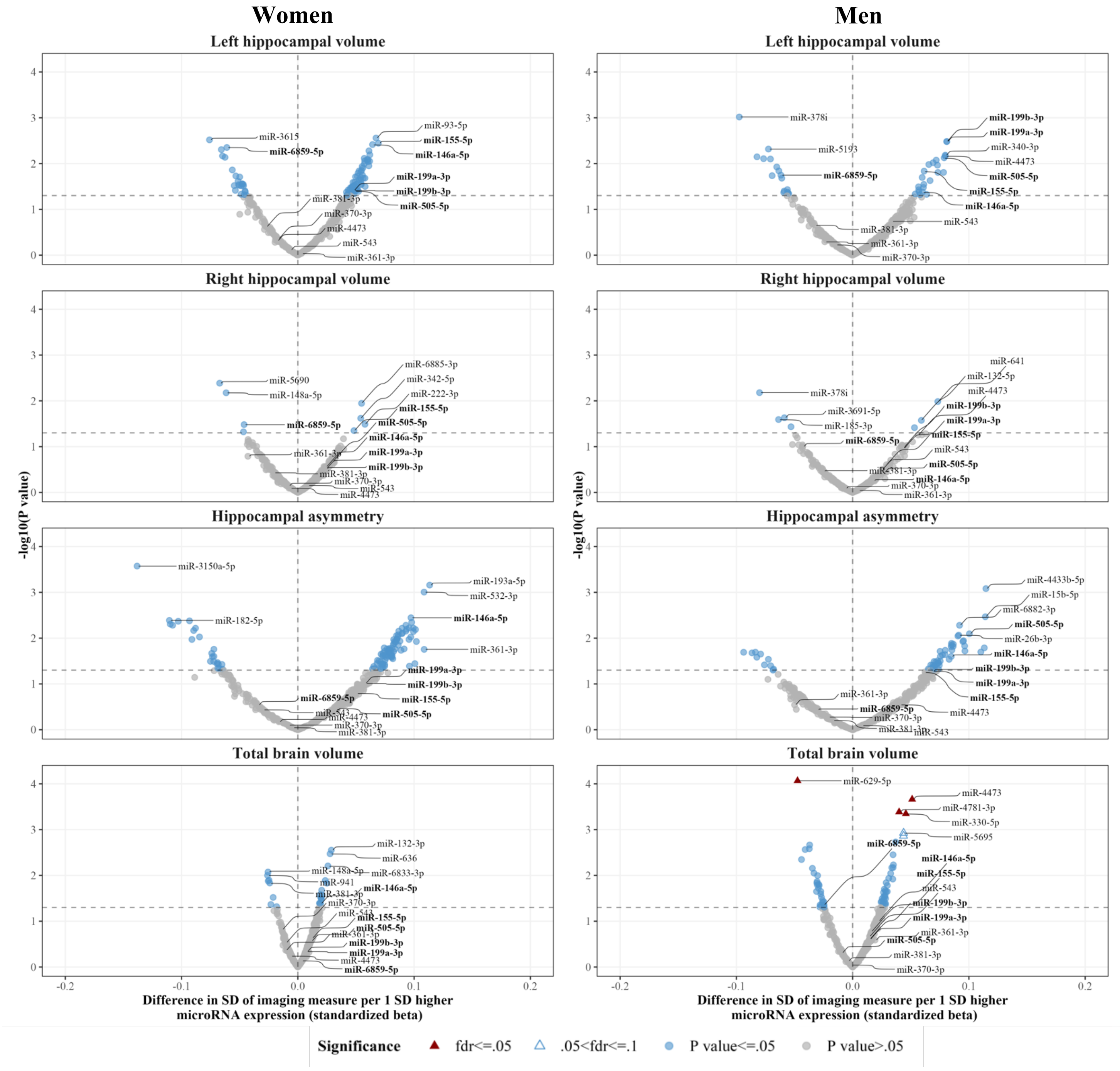


**Supplementary Figure S6:** Sex stratification, cross-sectional analysis
The volcano plots show the association of microRNAs with brain imaging measures in women and men. MicroRNAs significantly associated with HCV in the main analysis which included all participants have been annotated with bold letters.
Abbreviations: HCV. Hippocampal Volume; SD, Standard Deviation; fdr, False Discovery Rate

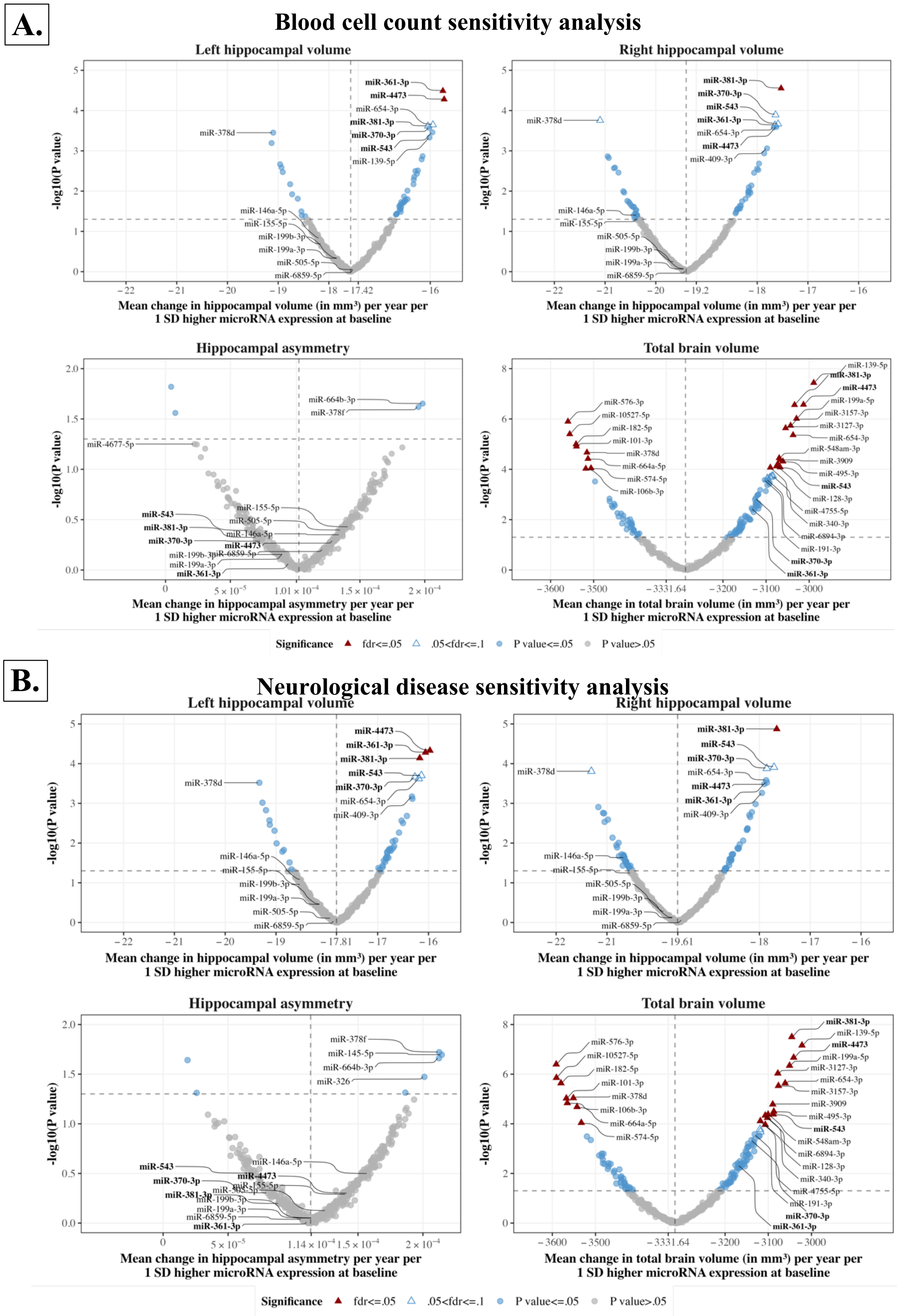


**Supplementary Figure S7:** Sensitivity analyses of the longitudinal association of microRNAs with brain imaging measures.
The volcano plots show the association of baseline microRNA expression with the rate of change of brain imaging measures when adjusting for baseline blood cell counts in addition to age, sex and technical variables (**A.**) and after removing participants diagnosed with dementia, Parkinson's disease, multiple sclerosis and hippocampal sclerosis (**B.**). MicroRNAs annotated with bold letters were significantly associated with hippocampal volume change across time in the main analysis. To facilitate plotting, different axis scales have been used for hippocampal volume change, asymmetry change, and total brain volume change.
Abbreviations: SD, Standard Deviation; fdr, false discovery rate

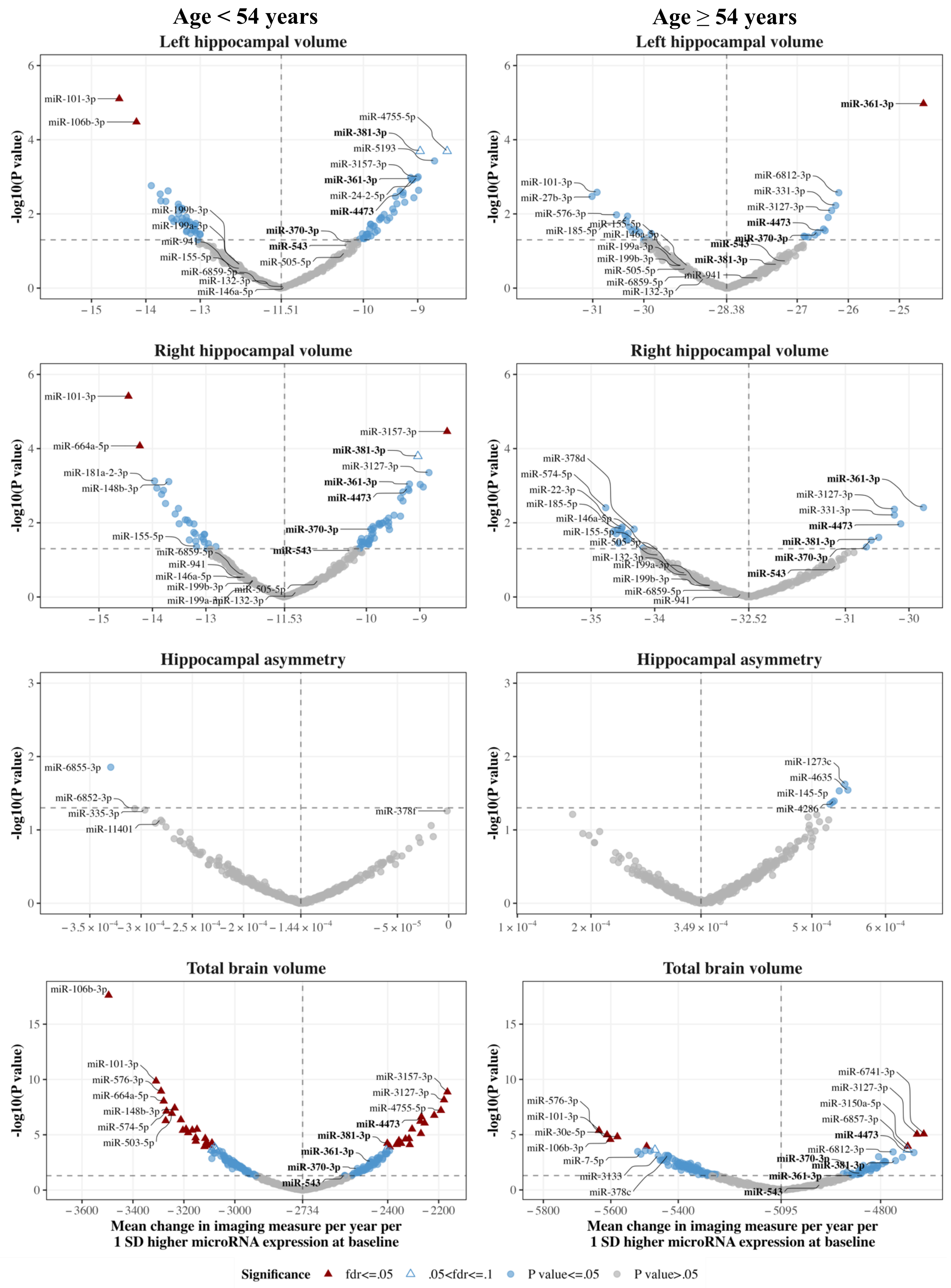


**Supplementary Figure S8:** Age stratification, longitudinal analysis

The volcano plots show the association of baseline microRNA expression with the yearly change of brain imaging measures in younger (age < 54 years at baseline) and older (age ≥ 54 years at baseline) participants. MicroRNAs significantly associated with hippocampal atrophy rate in the main analysis have been annotated with bold letters. Please note that, to facilitate plotting, different axis scales have been used.

Abbreviations: SD, Standard Deviation; fdr, False Discovery Rate

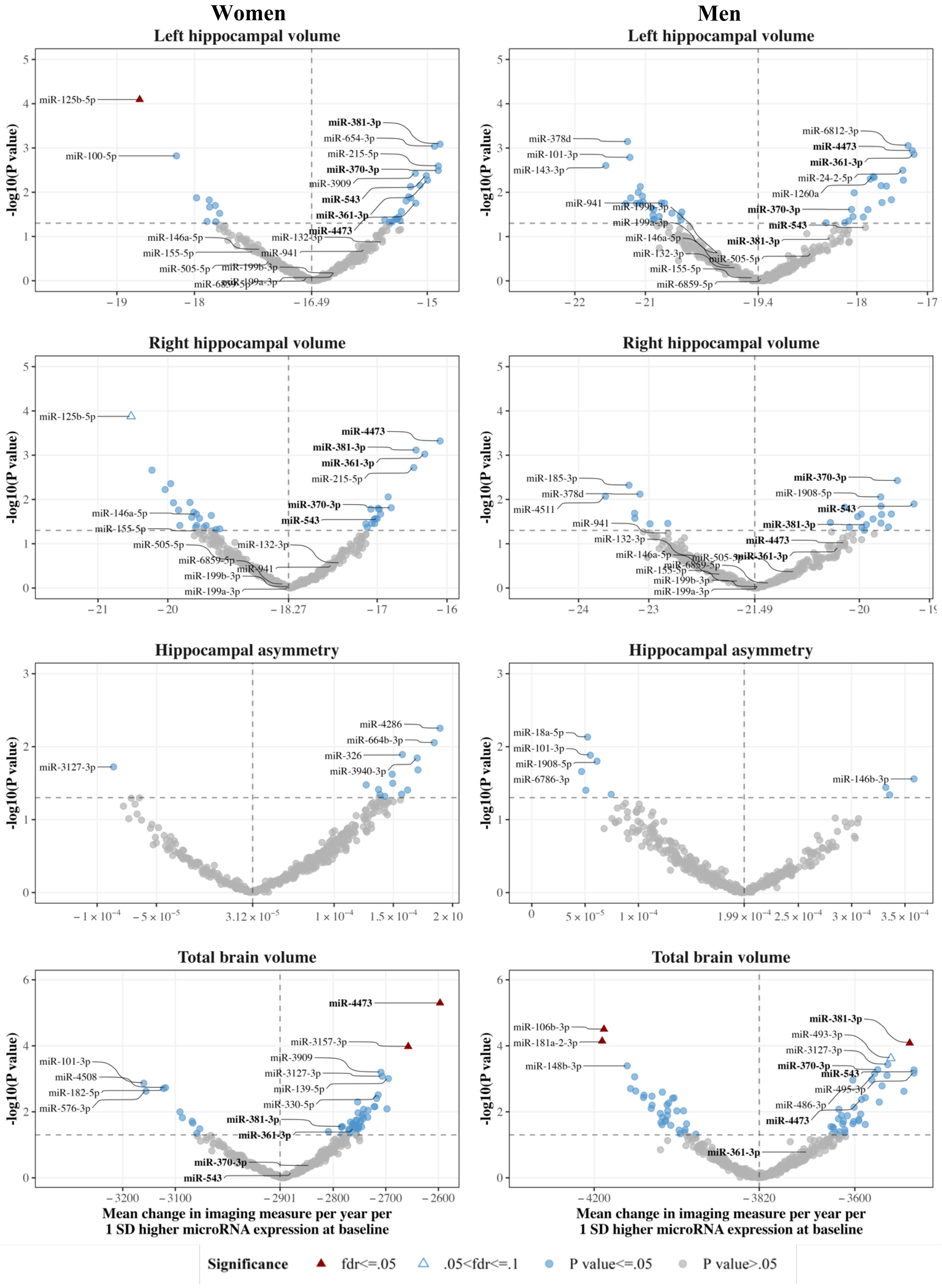


**Supplementary Figure S9:** Sex stratification, longitudinal analysis
The volcano plots show the association of baseline microRNA expression with the yearly change of brain imaging measures in women and men. MicroRNAs significantly associated with hippocampal atrophy rate in the main analysis have been annotated with bold letters. Please note that, to facilitate plotting, different axis scales have been used.
Abbreviations: SD, Standard Deviation; fdr, False Discovery Rate

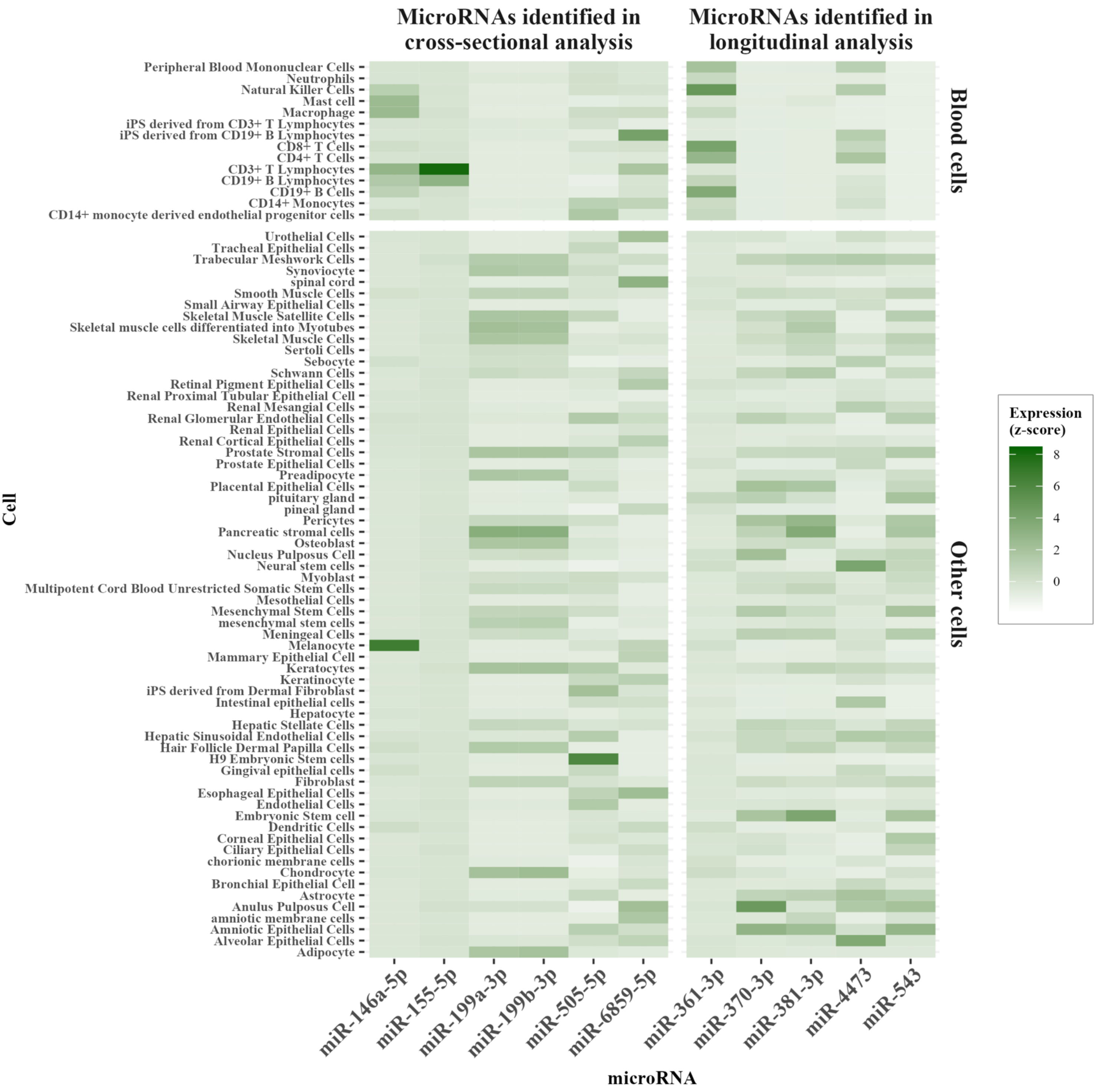


**Supplementary Figure S10:** MicroRNA expression in cells.
The heatmap shows relative miRNA expression (converted to a z-score for each miRNA) in various cell lines, for the microRNA associated with hippocampal volume cross-sectionally or longitudinally.

**A.** **Number of target genes identified in functional genomics analysis**

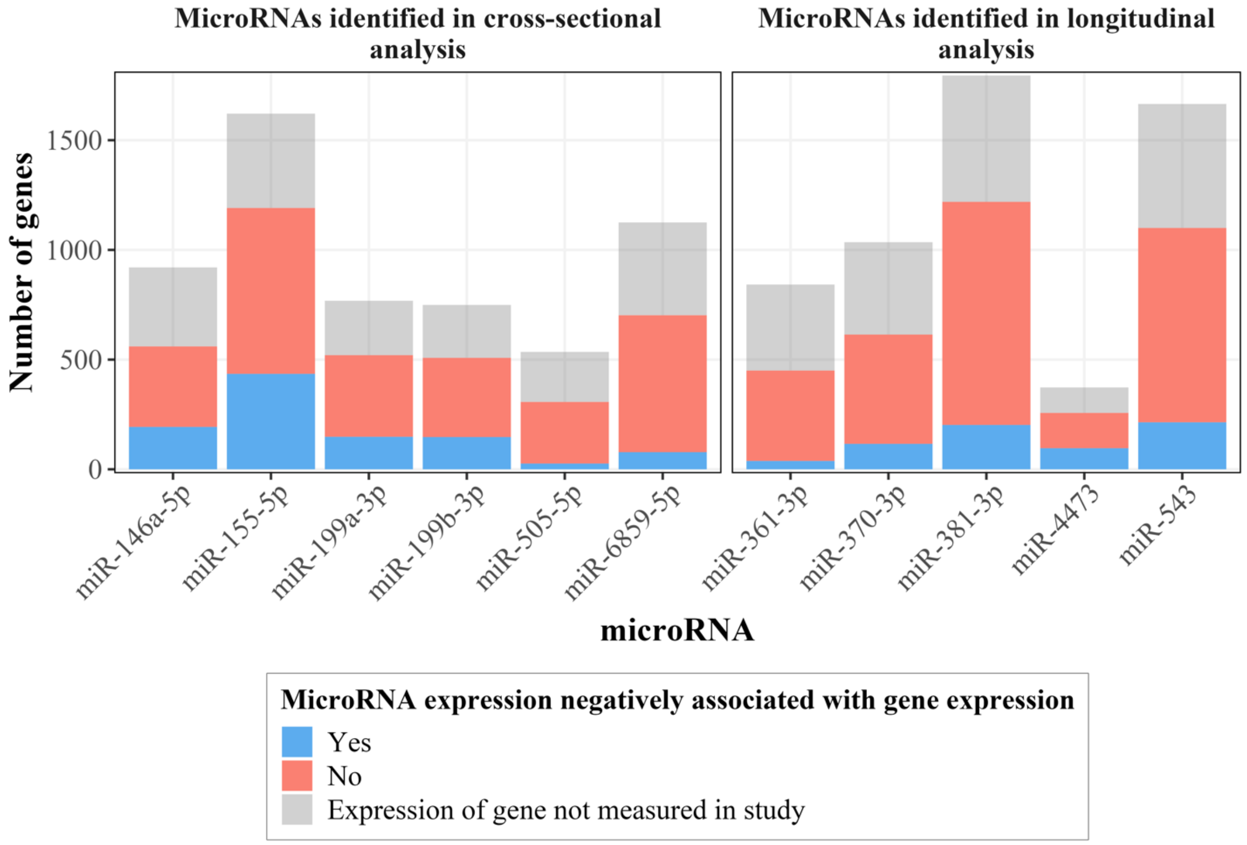


**B.** **Overlap of target genes identified in functional genomics analysis**

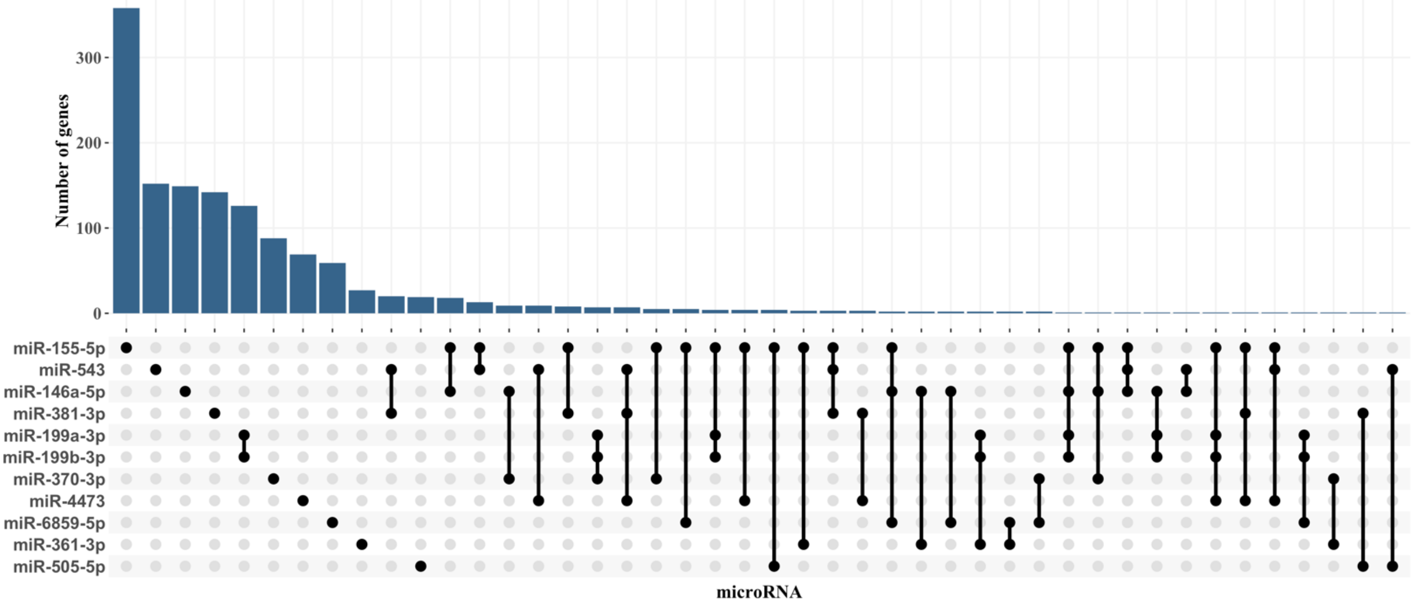


**Supplementary Figure S11:** MicroRNA target genes identified through functional genomics
The bar plot (**A.**) shows the number of genes that were computationally predicted to be targeted by hippocampus-related microRNAs, and the number of these predicted targets for which microRNA expression was negatively associated with gene expression. The association of microRNAs with some predicted target genes could not be examined, as expression of these genes was not measured in our study or was filtered out during quality control. These genes are shown with gray color. The upset plot (**B.**) shows the number and overlap of target genes that each microRNA was negatively associated with.

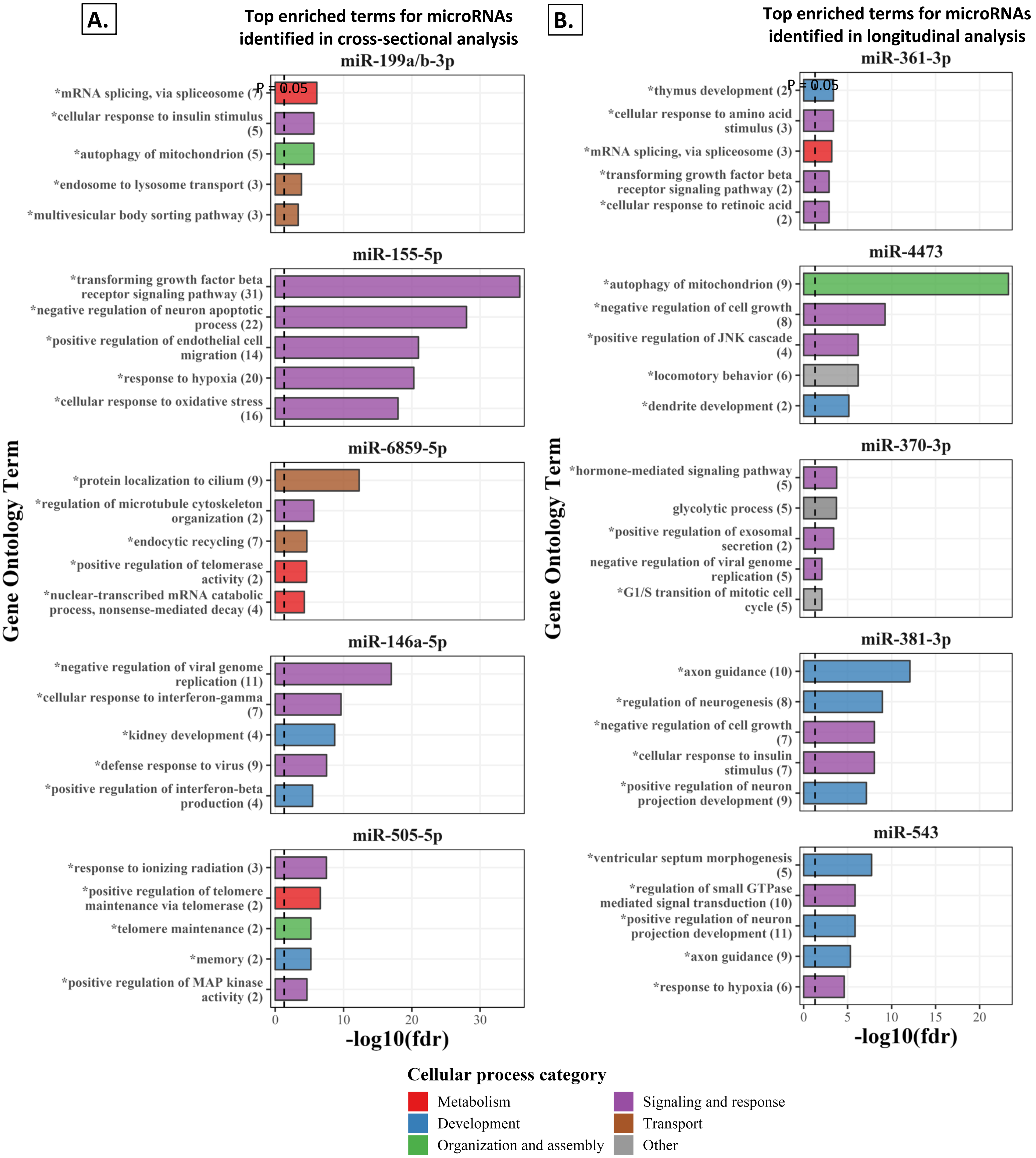


**Supplementary Figure S12:** Enrichment analysis of microRNA target genes expressed in the hippocampus. Results of *Gene ontology: Biological Process* enrichment analysis of microRNA target genes, filtering for genes with high expression in the hippocampus. The top 5 significantly enriched pathways (lowest p-value) are shown in the bar plots, for each microRNA associated with **A.** hippocampal volume cross-sectionally and **B.** hippocampal volume change rate longitudinally. Pathways marked with an asterisk **(*)** were enriched among target genes in the main analysis, without accounting for hippocampal expression. Significantly enriched pathways for each microRNA were grouped in broad categories, indicated by bar colors. Numbers in parentheses next to microRNA names indicate the total number of target genes of this microRNA that was included in *Gene Ontology.* P-values have been adjusted for multiple testing using the Benjamini-Hochberg false discovery rate (fdr) method.

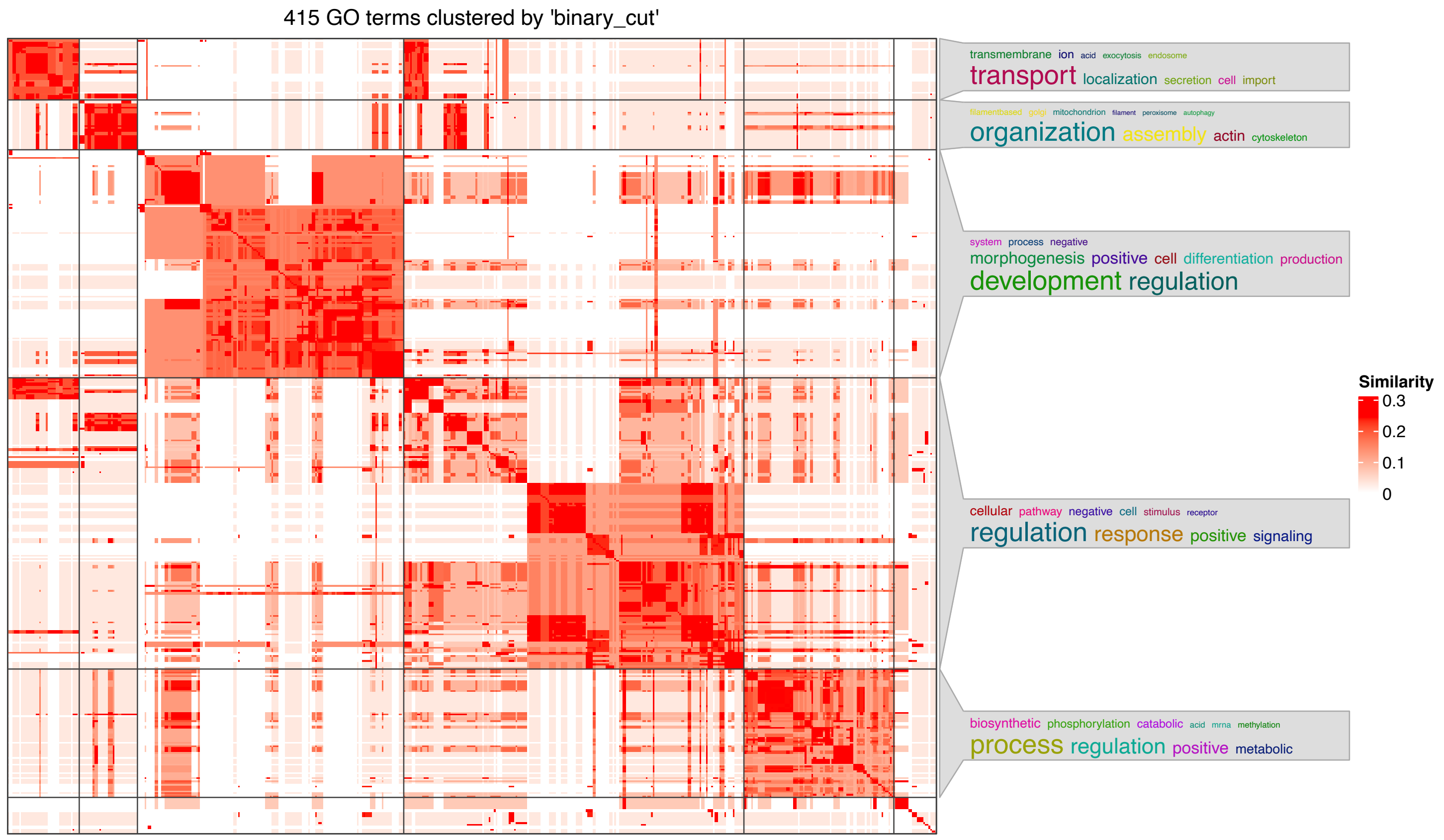


**Supplementary Figure S13:** Clustering of *Gene Ontology: Biological Processes* with the *SimplifyEnrichment* package. The heatmap shows the similarity matrix of terms, determined based on gene overlap. The word clouds show the most commonly appearing terms in each cluster, and were used to assign cluster names.
